# Brain-Controlled Epidural Spinal Stimulation for Upper-Limb Motor Function after Tetraplegia

**DOI:** 10.1101/2025.07.02.25330433

**Authors:** Atsushi Sasaki, Rizaldi Ahmad Fadli, Akiko Yuasa, Zachary Boogaart, Hiroki Saito, Nevena Musikic, Roberto de Freitas, Nicolo Macellari, Kevin C. Davis, Nilanjana Datta, Guilherme S. Piedade, Seth Tigchelaar, Michael E. Ivan, W. Dalton Dietrich, Naaz Desai, Vera Zivanovic, Milos R. Popovic, Elvira Pirondini, Jonathan Jagid, Abhishek Prasad, James D. Guest, Marco Capogrosso, Joacir Graciolli Cordeiro, Matija Milosevic

**Affiliations:** The Miami Project to Cure Paralysis, University of Miami Miller School of Medicine, FL, USA; Department of Neurological Surgery, University of Miami Miller School of Medicine, FL, USA; Graduate School of Arts and Sciences, The University of Tokyo, Tokyo, Japan; Department of Biomedical Engineering, University of Miami, FL, USA; Faculty of Medicine, Fujita Health University, Aichi, Japan; Japan Society for the Promotion of Science, Tokyo, Japan; Medical Scientist Training Program, University of Miami Miller School of Medicine, FL, USA; Department of Rehabilitation, Tokyo University of Technology, Tokyo, Japan; Center for Human Movement, Tokyo University of Technology, Tokyo, Japan; Rehab Neural Engineering Labs, University of Pittsburgh, Pittsburgh, PA, USA; Department of Neurological Surgery, University of Pittsburgh, Pittsburgh, PA, USA; KITE Research Institute – University Health Network, Toronto, ON, Canada; Institute of Biomedical Engineering, University of Toronto, Toronto, ON, Canada; CRANIA, University Health Network, Toronto, ON, Canada; Department of Bioengineering, University of Pittsburgh, Pittsburgh, PA, USA

## Abstract

Spinal cord injury (SCI) disrupts descending motor pathways, leaving individuals with tetraplegia reliant on residual neural connections for voluntary movement and limited recovery despite extensive rehabilitation. Epidural spinal cord stimulation (ESCS) has emerged as a promising neuromodulation strategy that can amplify spinal sensorimotor pathways, enabling residual circuits to respond more effectively to attempted voluntary commands. However, most approaches deliver tonic stimulation rather than in response to volitional intention, limiting the integration of cortical commands and spinal activation that may enhance both neuroprosthetic utility and the potential for recovery. Here, we present an implantable brain-computer interface (BCI) that decodes attempted movement from electrocorticography signals to trigger cervical ESCS during upper-limb motor tasks in an individual with chronic, motor-complete cervical SCI. We demonstrated that both BCI-driven and tonic ESCS immediately enhanced motor function compared to no stimulation, with BCI-ESCS producing greater improvements in grip force and reaching accuracy. Parallel to these assistive effects, BCI-ESCS was associated with changes in corticospinal and spinal excitability after a single session, whereas no comparable changes were observed following tonic stimulation. Four weeks of BCI-ESCS use further resulted in meaningful improvements in hand motor function exceeding clinical improvement thresholds, with selected gains persisting at one-month follow-up. Together, these findings establish a translational proof-of- concept for implantable BCI-ESCS and provide preliminary evidence for the feasibility of intention-driven neuromodulation to support both neuroprosthetic assistance and sustained improvements in upper-limb motor function in chronic motor-complete tetraplegia.

**One-Sentence Summary:** Implanted brain-controlled spinal stimulation provides neuroprosthetic assistance and supports motor recovery in tetraplegia.

## INTRODUCTION

Spinal cord injury (SCI) is a life-altering condition that disrupts descending motor pathways, leading to severe impairments in voluntary motor control. Among individuals with cervical SCI, tetraplegia substantially limits upper-limb function, reducing independence and quality of life (*1*). Approximately two-thirds of SCIs occur at the cervical level (63.4% at C1–C7), often resulting in tetraplegia and impairing upper-limb control (*2*). Although individuals with incomplete injuries (American Spinal Injury Association Impairment Scale (AIS) B-D) retain some degree of sensorimotor function, those with complete injuries (AIS A) typically exhibit low to no voluntary movement below the lesion level with minimal recovery potential (*3*, *4*). Despite extensive rehabilitation efforts, standard physical and occupational therapy often leads to limited motor recovery, necessitating novel neuromodulatory strategies to interface with remaining sensorimotor circuits to augment task-relevant upper-limb motor output.

Epidural spinal cord stimulation (ESCS) has emerged as a promising tool for increasing the excitability of spinal sensorimotor circuits and enabling these circuits to respond more effectively to residual or attempted voluntary commands. The potential of ESCS to augment upper-limb function was explored in non-human primates, where cervical ESCS was found to enhance voluntary reaching and grasping by modulating spinal motoneurons (*5*, *6*). In humans, early studies demonstrated that ESCS can facilitate volitional lower-limb movement in individuals with chronic paraplegia (*7–11*). Lu et al. (*12*) further provided human evidence that cervical ESCS can enhance voluntary hand function in individuals with chronic tetraplegia by engaging spinal circuits. More recently, tonic cervical ESCS has been shown to improve grip strength and movement kinematics in individuals with post-stroke hemiparesis (*13*, *14*). Although these studies highlight the immediate assistive benefits of cervical ESCS, most existing approaches deliver tonic stimulation, rather than using the user’s own neural activity to initiate stimulation when attempted movements are detected.

Brain-computer interface (BCI) technology enables conversion of residual neural commands during attempted movements into control signals for external devices by decoding motor intention and providing real-time stimulation to enable upper-limb muscle activation (*15–17*). Unlike conventional tonic ESCS, BCI-controlled neuromodulation has the potential to drive spinal stimulation according to the user’s own volitional neural activity (*18*, *19*). This creates a human-in-the-loop control architecture in which intended motor commands are detected in real time and translated into targeted activation of spinal circuits. Preclinical studies demonstrated that BCI-controlled neuromodulation can induce adaptive cortical changes and immediate neuroprosthetic effects in animal models (*6*, *20*, *21*). Lorach et al. (*19*) recently demonstrated that an implantable brain-spinal interface can restore lower-limb function in an individual with incomplete SCI, establishing the clinical feasibility of linking cortical signals to spinal stimulation for locomotor control. However, the clinical translation of an implantable brain-spinal interface for upper-limb function, particularly in individuals with complete cervical SCI, has not yet been realized. This closed-loop application requires feasibility testing, including detection of attempted movement from neural activity, task-specific configuration of cervical stimulation, and stimulation timing that supports movement attempts while minimizing activation at rest.

Here, we present a first-in-human implantable electrocorticography (ECoG)-based BCI coupled with cervical ESCS for intention-driven upper-limb neuromodulation in an individual with chronic motor-complete cervical SCI (AIS A). We tested whether cortical activity could be used to trigger task-specific ESCS during attempted upper-limb movements, and if this BCI-ESCS system could produce immediate motor assistance, facilitate motor performance, and enhance neurophysiological readouts of corticospinal and spinal excitability. Our primary objectives were as follows. First, we assessed the immediate assistive effects of BCI-ESCS and tonic ESCS on grip strength, reaching kinematics, and hand function. Second, we evaluated the single-session effects of BCI-ESCS and tonic ESCS on corticospinal and spinal reflex circuit excitability. Third, we determined the longitudinal effects of four-week BCI-ESCS use on upper-limb motor performance and neurophysiological readouts. By integrating ECoG-based motor-intention detection with epidural spinal stimulation, this study investigates the feasibility of an implantable brain-controlled cervical spinal stimulation system for upper-limb motor assistance after chronic complete SCI.

## RESULTS

### Study design and experimental setup

This study implemented an implantable BCI-controlled ESCS approach in an individual with chronic motor-complete cervical SCI. The participant was a male in his 20’s with a chronic (11 years post-injury) motor-complete AIS A injury at the C4–C5 neurological level, confirmed by an International Standards for Neurological Classification of Spinal Cord Injury (ISNCSCI) examination (see “Clinical assessments” section of Supplementary Materials) performed immediately prior to study initiation. This classification was further supported by magnetic resonance imaging (MRI) (Fig. 1), which showed a focal lesion at the C4 level, and by the absence of measurable motor evoked potentials (MEPs) and voluntary electromyographic (EMG) activity at baseline in intrinsic hand muscles innervated below the C5 level (*22*), including the first dorsal interosseous (FDI), abductor digiti minimi (ADM), and abductor pollicis brevis (APB) (Fig. S1). Details of EMG acquisition are provided in the “Electromyography acquisition system” section of the Supplementary Materials. The participant had undergone anterior cervical discectomy and fusion (C3–C5) and subsequently received an ECoG implant over the right-hand knob area of the motor cortex as part of another clinical trial evaluating BCI utility following tetraplegia (*23*, *24*), before enrolling in the current study where percutaneous leads were implanted to test the feasibility of BCI-driven cervical ESCS over four weeks (NCT06533969).

**Fig. 1.**
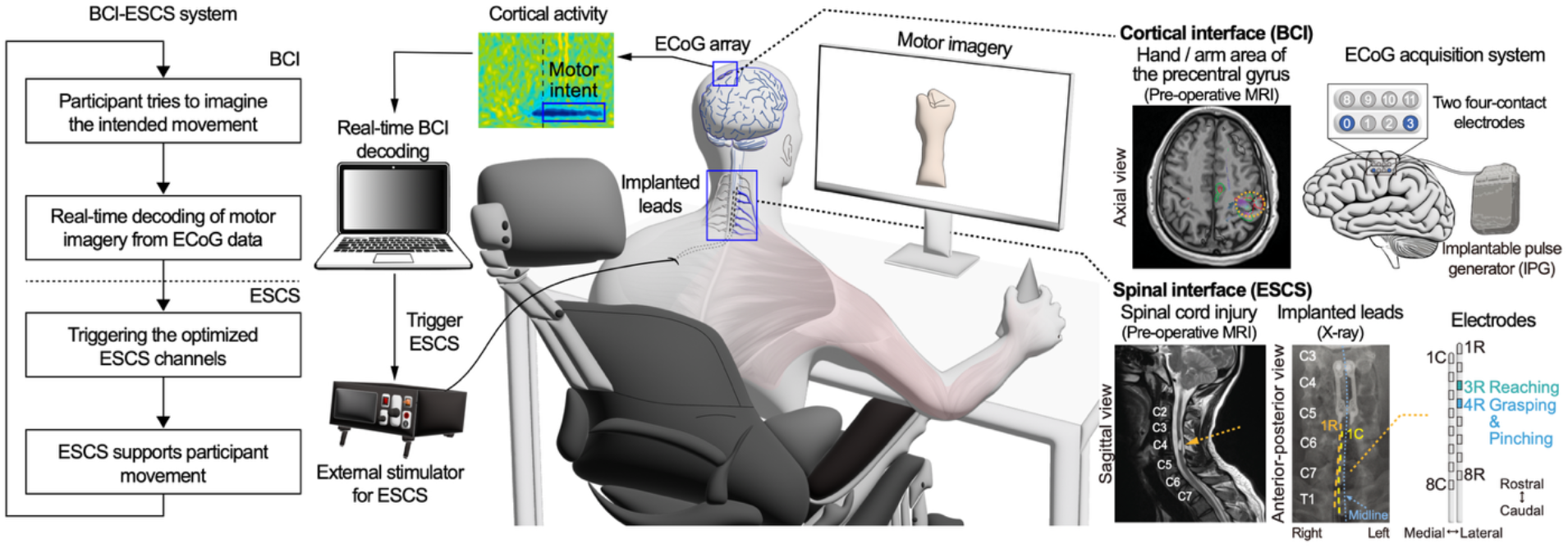
Implantable BCI-ESCS system for upper-limb motor function following chronic SCI. The brain-computer interface (BCI)–epidural spinal cord stimulation (ESCS) system integrates attempted movement-related cortical activity detection with cervical spinal stimulation to support upper-limb motor output in an individual with chronic complete cervical spinal cord injury (C4–C5; AIS A). The participant attempts to imagine a movement, which is detected through an implanted electrocorticography (ECoG) array placed over the precentral gyrus, as identified in preoperative magnetic resonance imaging (MRI) (*23*). ECoG signals were acquired using an implantable pulse generator (IPG), which was also used to deliver single-pulse motor cortex stimulation for motor evoked potential (MEP) measurements. The BCI system decodes cortical activity in real time and, upon detection of motor intention, triggers an external stimulator to deliver optimized ESCS via percutaneously implanted epidural leads. The spinal X-ray image confirms the placement of epidural electrodes over the C5–T1 vertebral levels, with specific channels targeting functional tasks such as reaching (contact: 3R) and grasping/pinching (contact: 4R). This approach is designed to initiate cervical ESCS following attempted movements, providing a brain-controlled platform for task-specific upper-limb neuromodulation.

In the current study, BCI-ESCS was implemented by detecting attempted movement-related cortical activity to initiate cervical ESCS during motor tasks. Motor imagery was decoded using a one-degree-of-freedom classifier (*25–29*) by monitoring event-related desynchronization (ERD) activity from ECoG recordings (Ch 0-3) within the 8–14 Hz band. These channels and frequency range were identified during a calibration session based on the strongest ERD during upper-limb motor imagery tasks (see the “BCI system” section). During task performance, the BCI detected motor imagery when the cortical signal power remained below a threshold for one second within a 15-second window after the Go cue (*25–29*), which triggered ESCS. The decoding success rate was 83.8±4.2 %. ESCS was delivered via task-specific contacts selected from two percutaneous eight-contact leads implanted in the dorsal epidural space from vertebral levels C5–T1 caudal to the injury site (Fig. 1; “Surgical procedure” section of the Supplementary Materials). The leads were positioned unilaterally on the right side over the dorsal roots under fluoroscopic guidance to selectively activate right upper-limb motor pools (*13*), as also confirmed by post-activation depression in ESCS-evoked posterior root muscle (PRM) reflexes (Fig. S2) (*30*, *31*). For each task, contacts were selected to preferentially recruit the relevant upper-limb muscles, with task-specific configurations varying across sessions (Fig. S3; see the "Epidural spinal cord stimulation (ESCS) system" section). Stimulation was delivered at 80 Hz using a 200-µs biphasic waveform, with intensities selected to facilitate volitional movement without eliciting involuntary muscle contraction at rest (1.0–3.4 mA), which were near or just below the motor threshold to elicit PRM reflex responses (*8*, *13*).

BCI-ESCS effects were evaluated through three distinct analyses: (1) Assistive effects - Immediate functional effects of BCI-ESCS compared to no-stimulation and tonic ESCS conditions; (2) Single-session effects - Changes in neurophysiological outcomes after a single BCI-ESCS session compared with tonic stimulation; and (3) Longitudinal effects - Functional and neurophysiological outcomes following four weeks of BCI-ESCS use. A total of 19 sessions were completed over four weeks following ESCS lead implantation: 16 sessions were BCI-ESCS and 3 were tonic ESCS, during which tonic stimulation was applied throughout the session.

### Assistive effects of BCI-ESCS and tonic ESCS on grip strength, reaching, and hand motor function

To assess immediate assistive effects, we compared no-stimulation, Tonic-ESCS (stimulation delivered throughout the session), and BCI-ESCS (stimulation triggered by motor intention) using identical stimulation contacts, intensities, and frequency (Fig. 2). During a maximum voluntary contraction (MVC) of the gripping task (Fig. 2A; Table S1; “Maximum grip assessment” section of Supplementary Materials), both stimulation modes immediately enhanced grip force and volitional EMG activation in the flexor carpi radialis (FCR) and extensor carpi radialis (ECR) muscles in an individual who presented with no detectable voluntary upper-limb EMG activity during the no-stimulation condition. These findings demonstrate that cervical ESCS can augment motor output, likely by increasing the responsiveness of spinal circuits to residual descending commands (*13*, *14*). Notably, BCI-ESCS produced higher grip force and EMG activity, suggesting that coupling stimulation to the user’s motor intention may generate stronger stimulation-assisted grip output than tonic stimulation alone.

**Fig. 2.**
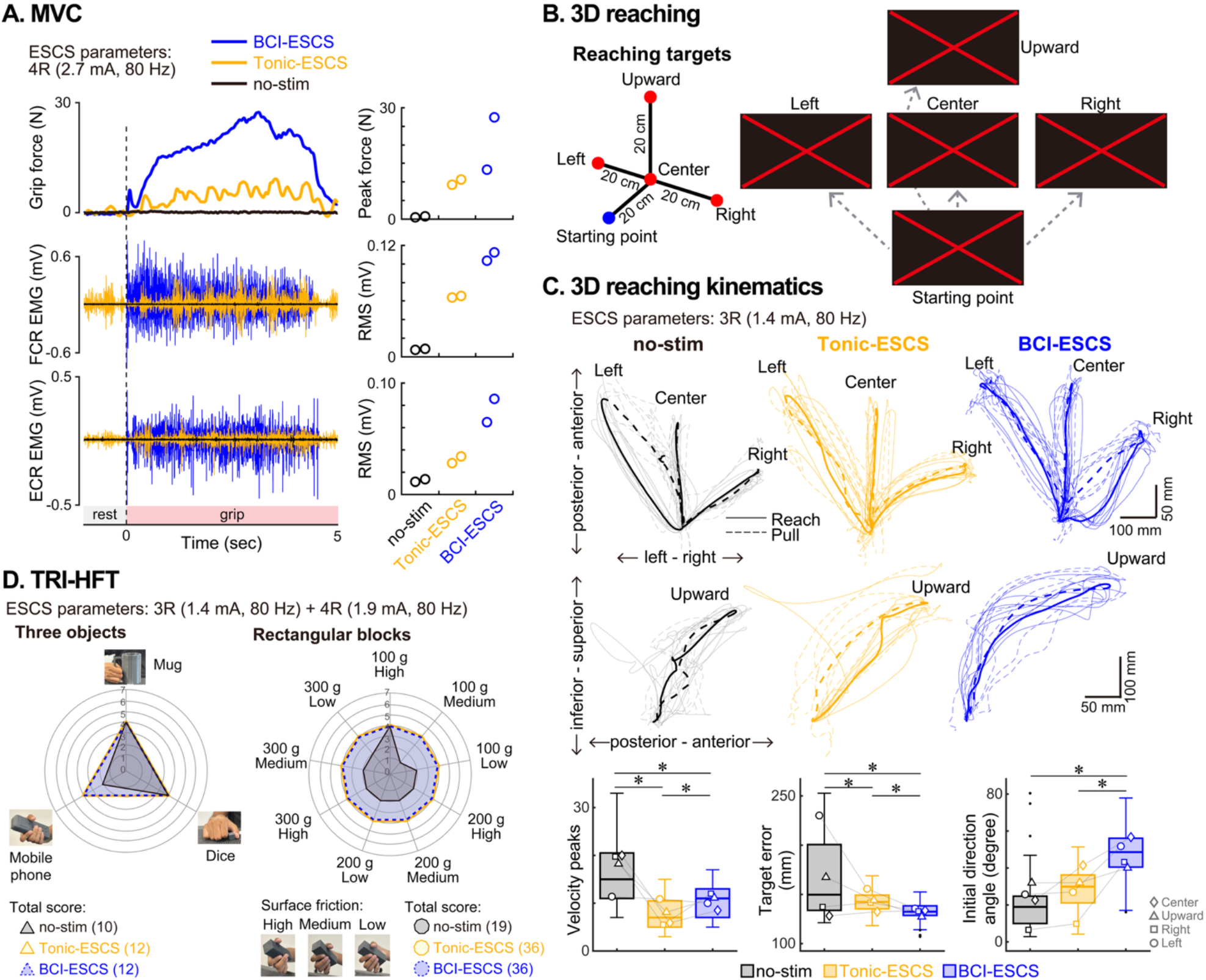
BCI-ESCS and Tonic-ESCS immediately assist grip force and upper-limb motor performance. **(A)** Grip force and EMG activity from the flexor carpi radialis (FCR) and extensor carpi radialis (ECR) muscles during a 5-second maximal voluntary contraction (MVC), with two trials per condition. Representative traces are shown for no-stimulation (black), Tonic-ESCS (yellow), and BCI-ESCS (blue). Both stimulation conditions resulted in higher peak grip force and EMG root-mean-square (RMS), with greatest facilitation observed during BCI-ESCS. **(B)** 3D reaching task. The participant performed reaching movements toward four target positions (left, center, right, and upward) from a fixed starting position as accurately as possible, with eight repetitions per target. **(C)** Reaching kinematics under no-stimulation (black), Tonic-ESCS (yellow), and BCI-ESCS (blue). Representative trajectories are shown for each target direction. Boxplots show pooled kinematic metrics across four target directions (8 attempts × 4 targets per condition), including velocity peaks (movement smoothness), target error (endpoint accuracy), and initial direction angle (deviation from the straight-line trajectory). In each boxplot, the central line indicates the median, the box indicates the interquartile range (IQR), whiskers extend to 1.5×IQR, dots represent outliers, and asterisks indicate statistical significance based on bootstrap analysis (n=10,000 resamples). Different symbols indicate the mean for each target. Both Tonic-ESCS and BCI-ESCS showed fewer velocity peaks and reduced target error compared to no-stimulation. Tonic-ESCS showed the fewest velocity peaks, whereas BCI-ESCS showed the lowest target error and the largest initial direction angle. **(D)** Functional assessments using the Toronto Rehabilitation Institute-Hand Function Test (TRI-HFT). Radar plots show scores for each individual object in the three-object manipulation and rectangular block tasks during no-stimulation (black), Tonic-ESCS (yellow), and BCI-ESCS (blue) conditions. Each spoke represents an individual object or task, and greater radial distance from the center indicates better performance. Total scores are shown below each radar plot. Both BCI-ESCS and Tonic-ESCS improved task performance compared to no stimulation, with the mobile phone score increasing from 2 to 4 points and the total rectangular block score increasing from 19 to 36 points. **NOTE**: All assistive-effect assessments were compared using identical ESCS settings (contacts, stimulation amplitude, and frequency) for the BCI and Tonic modes.

Beyond hand force, we also evaluated upper-limb coordination during a 3D reaching task toward four targets (left, right, center, and upward) (Fig. 2B; “3D reaching assessment” of Supplementary Materials). Compared to no-stimulation, both BCI-ESCS and Tonic-ESCS improved movement smoothness, as reflected by fewer velocity peaks (Fig. 2C; Table S1) and endpoint accuracy, as reflected by smaller target error (Fig. 2C; Table S1), further supporting the assistive benefits of cervical ESCS. Specifically, BCI-ESCS achieved the lowest endpoint error, whereas Tonic-ESCS produced smoother trajectories (Fig. 2C; Table S1), reflecting task-dependent differences between stimulation modes. BCI-ESCS generated a larger deviation at movement onset (initial direction angle), likely reflecting altered movement initiation during intention-triggered stimulation. Muscle synergy analysis from 16 upper-limb muscles (shoulder to forearm) during reaching further illustrates that both stimulation modes can alter neuromuscular coordination as reflected by a greater number of muscle modules compared to no stimulation, suggesting a shift from broad co-activation toward more functionally distinct activation patterns (*32*, *33*) (Fig. S4).

Improvements in grip and reaching performance also translated to immediate gains in functional object manipulation, with scores on the Toronto Rehabilitation Institute-Hand Function Test (TRI-HFT) improving under BCI-ESCS and Tonic-ESCS conditions (Fig. 2D). In the mobile phone task, both stimulation conditions improved the score from 2 to 4 points compared with no stimulation. Most notably, rectangular block scores increased from 19 to 36 points under both stimulation modes, highlighting the substantial contribution of cervical ESCS on motor functional gains.

### Single-session effects of BCI-ESCS and Tonic-ESCS on corticospinal and spinal excitability

We examined the short-term neurophysiological and functional effects of BCI-ESCS and Tonic-ESCS approaches using a counterbalanced crossover design with two repeated sessions per condition (Fig. 3A). Each session consisted of 30 min of upper-limb motor tasks guided by a physical therapist, who followed an identical protocol across conditions. Motor tasks focused on reaching, grasping, and pinching with task-specific ESCS parameters for each session (Fig. S3). We assessed corticospinal excitability using motor evoked potentials (MEPs) elicited by single-pulse motor cortex stimulation. Spinal excitability was assessed using posterior root muscle (PRM) reflexes elicited by paired-pulse ESCS, in which suppression of the second response confirmed the reflex origin of the evoked response (*30*, *31*). Maximum M-wave (M_max_) responses were also assessed to normalize MEPs and PRM reflex amplitudes and help reduce some of the influence of variability in peripheral recordings across conditions (*34*). All measures were obtained before (Pre) and immediately after (Post) each session. Alongside these primary neurophysiological outcomes, motor performance was assessed using the nine-hole peg test (9HPT) and box and block test (BBT) without stimulation (“Clinical assessments” section of Supplementary Materials).

**Fig. 3.**
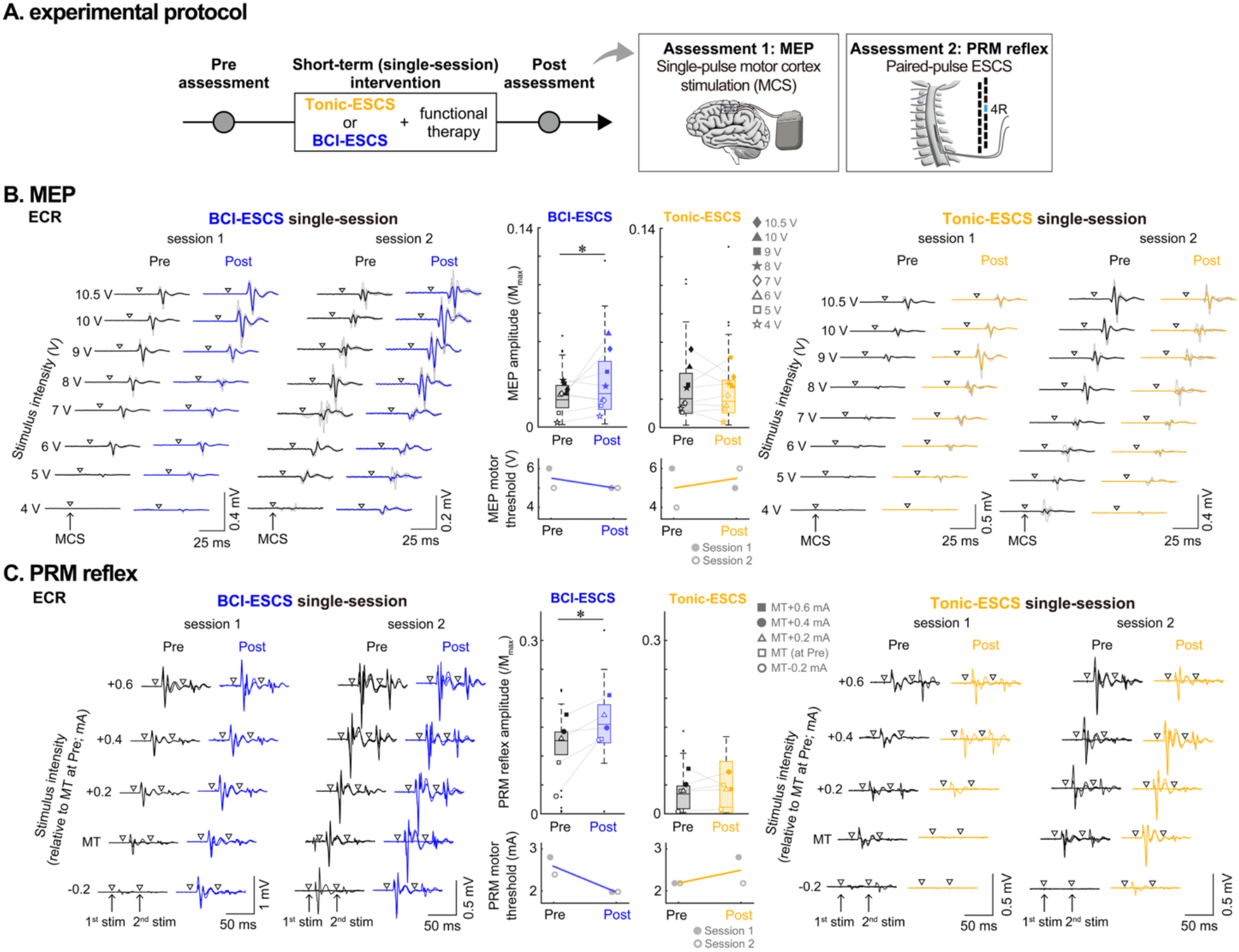
Corticospinal and spinal excitability following single-session BCI-ESCS and Tonic-ESCS use. **(A)** Experimental protocol. Single-session effects were assessed in a counterbalanced crossover design with two sessions per condition. Neurophysiological assessments were conducted before (Pre) and after (Post) each session in the extensor carpi radialis (ECR) muscle. **(B)** Corticospinal excitability measured via motor evoked potentials (MEPs) across 4–10.5 V stimulation intensities (8 intensities × 5 trials per session), with waveforms from both sessions shown for each BCI-ESCS (blue) and Tonic-ESCS (yellow) condition. Boxplots show that BCI-ESCS was associated with facilitation of MEP/M_max_ amplitudes, whereas Tonic-ESCS resulted in no changes. Line plots show mean MEP motor threshold across two sessions, defined as the lowest stimulation intensity eliciting >50 µV peak-to- peak responses, with lower values observed following BCI-ESCS use. **(C)** Spinal excitability assessed via posterior root muscle (PRM) reflexes elicited by paired-pulse ESCS across stimulation intensities, with waveforms from both sessions shown for each BCI-ESCS (blue) and Tonic-ESCS (yellow) condition. Stimulation intensities were referenced to the Pre-measurement PRM reflex motor threshold (MT), ranging from MT-0.2 to MT+0.6 mA (5 intensities × 3 trials per session). Suppression of the second response relative to the first confirms dorsal root targeting. Boxplots show that BCI-ESCS was followed by larger PRM/M_max_ amplitudes, whereas Tonic-ESCS showed limited changes. Line plots show mean PRM reflex motor threshold across the two sessions, with lower values following BCI-ESCS use. **NOTE**: Across all boxplots, data are pooled across all stimulation intensities and sessions. The central line indicates the median, the box indicates interquartile range (IQR), whiskers extend to 1.5×IQR, and dots represent outliers. Different symbols indicate mean responses at each tested stimulation intensity. Asterisks indicate statistical significance based on bootstrap analysis (n=10,000 resamples).

BCI-ESCS use was associated with changes in corticospinal excitability, as evidenced by larger MEP/M_max_ amplitudes and lower MEP motor threshold in the ECR muscle (Fig. 3B; Table S2). Similarly, PRM/M_max_ amplitudes increased, and PRM reflex motor threshold decreased following BCI-ESCS (Fig. 3C; Table S2), indicating changes in spinal excitability. Consistent MEP and PRM reflex changes were also observed in the FCR muscle (Fig. S5; Table S2). In contrast, neurophysiological changes were not observed following the Tonic-ESCS sessions despite identical motor tasks and matched stimulation parameters (Fig. 3B-C; Fig. S5). Although these single-session findings should be interpreted cautiously given the single-participant design, the MEP and PRM responses provide complementary preliminary evidence that temporal coordination between motor intention and spinal activation may contribute to excitability changes across corticospinal and spinal circuits.

In addition to neurophysiological assessments, we evaluated short-term motor performance before and after each condition. The 9HPT remained unchanged in both conditions (0.0±0.0 to 0.0±0.0 pegs), and BBT scores changed from 4.0±0.0 to 6.0±0.0 and from 3.0±1.4 to 3.5±0.7 following Tonic-ESCS and BCI-ESCS, respectively. As the voluntary function scores remained at floor level and the observed changes fell below minimal detectable change thresholds (*35*, *36*), these assessments indicated no functional improvements following a single session in this individual with chronic motor-complete injury.

Overall, these findings suggest that BCI-driven stimulation may contribute to excitability changes across corticospinal and spinal circuits, prompting further investigation into whether repeated BCI-ESCS use over a four-week period could translate into functional gains.

### Longitudinal effects of BCI-ESCS use on voluntary hand function and corticospinal excitability

We investigated functional and neurophysiological changes following four weeks of BCI-ESCS use. Hand motor function during object manipulation was assessed alongside neurophysiological outcomes of corticospinal and spinal circuits, reaching kinematics, and ISNCSCI evaluation. Functional assessments were conducted at Baseline (the start of the longitudinal BCI-ESCS trial), Endpoint (the end of the trial), and Follow-up (one month after ESCS lead explantation), with neurophysiological assessments also conducted at three intermediate timepoints (Fig. 4A, Fig. S3). During BCI-ESCS use, the participant performed upper-limb motor tasks including reaching, grasping, pronation, supination, and pinching, guided by a physical therapist, lasting 30-40 min. Tasks progressed from basic movements to complex functional tasks, incorporating objects of varying size, weight, and texture to enhance motor engagement. As performance improved, tasks mimicking daily activities, such as drinking and eating, were incorporated to facilitate functional goals (see the “BCI-ESCS therapy protocol” section).

**Fig. 4.**
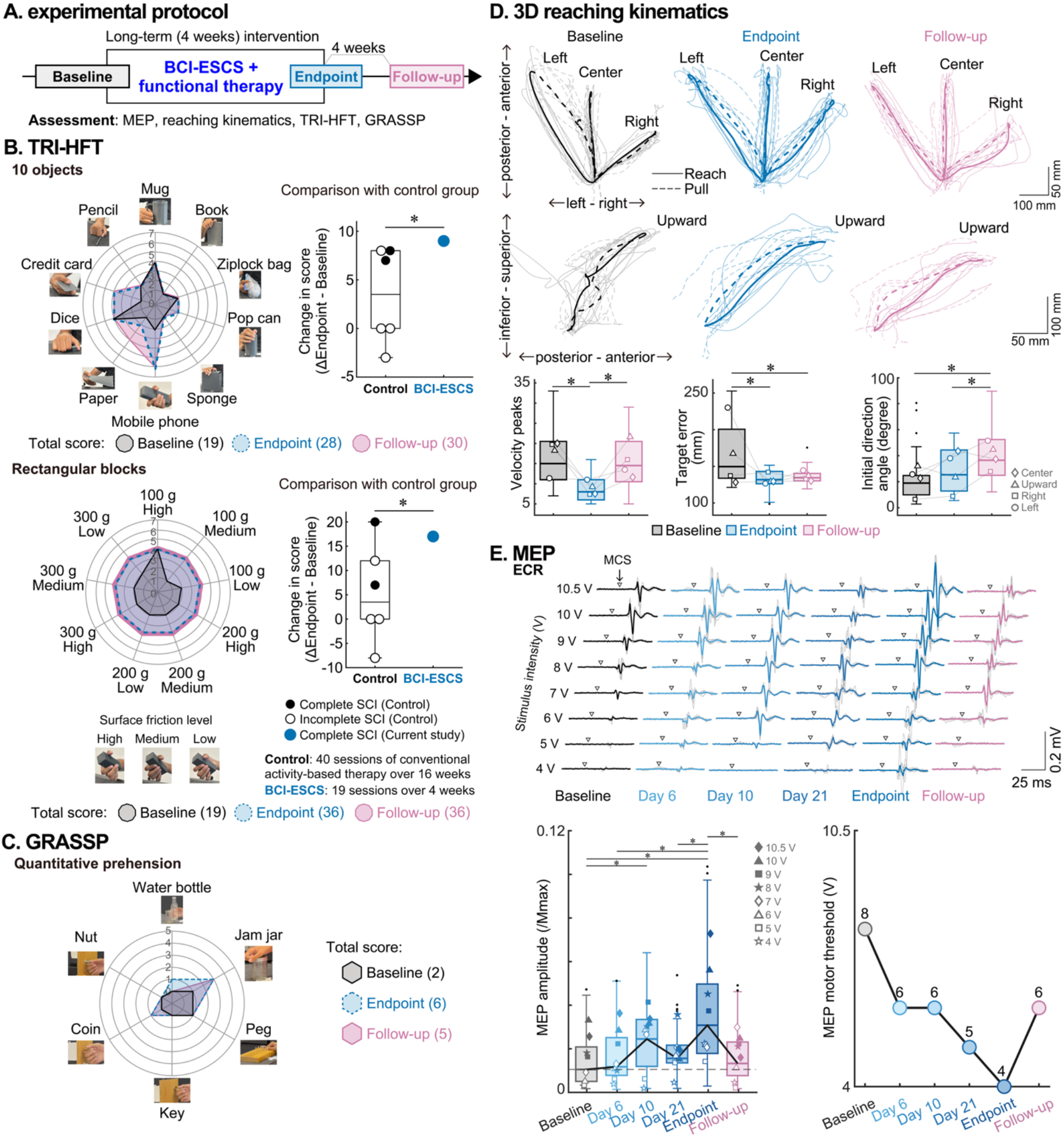
Functional performance and corticospinal excitability following long-term BCI-ESCS use. **(A)** Experimental protocol. Functional and clinical assessments were conducted at Baseline, at the start of the longitudinal BCI-ESCS intervention; Endpoint, at the end of the four-week BCI-ESCS use period; and Follow-up, 4 weeks after ESCS lead explantation, to evaluate the longitudinal effects of BCI-ESCS use (Fig. S3). **(B)** TRI-HFT performance. Radar plots show scores for each object in the 10-object manipulation and rectangular block tasks at Baseline (black), Endpoint (blue), and Follow-up (pink). Each spoke represents an individual object or task, and greater radial distance from the center indicates better performance. Total scores are shown below each radar plot. The 10-object score increased from 19 at Baseline to 28 at Endpoint and 30 at Follow-up, with notable gains in mobile phone, pop can, paper, and credit card manipulation. The rectangular block score increased from 19 at Baseline to 36 at Endpoint and remained at 36 at Follow-up. Boxplots compare the participant’s score changes (n=1) to those of a control cohort (n=2 with C5–C6 complete SCI and n=4 with C4–C7 incomplete SCI) who underwent conventional therapy only. **(C)** GRASSP quantitative prehension test. Radar plot shows scores for each individual task at Baseline (black), Endpoint (blue), and Follow-up (pink). The total score increased from 2 at Baseline to 6 at Endpoint and 5 at Follow-up, with notable gains in the water bottle, jam jar, and coin tasks. **(D)** Reaching kinematics. Reaching trajectories are shown for Baseline, Endpoint, and Follow-up assessments. Boxplots of velocity peaks, target error, and initial direction angle show pooled kinematic metrics across all four target directions (8 attempts × 4 targets per assessment timepoint). Velocity peaks were reduced at Endpoint but returned toward Baseline at Follow-up. Target error was reduced at Endpoint and remained lower at Follow-up. The initial direction angle increased at Follow-up, indicating altered initial trajectory formation. **(E)** Corticospinal excitability measured via motor evoked potentials (MEPs). MEP waveforms from the extensor carpi radialis (ECR) muscle are shown across all six assessment timepoints: Baseline (2 days before ESCS lead implantation), Day 6, Day 10, Day 21, Endpoint (Day 22), and Follow-up (4 weeks after ESCS lead explantation). The boxplot shows MEP/M_max_ amplitudes pooled across all tested stimulation intensities at each assessment timepoint (8 intensities × 5 trials per assessment timepoint). MEP/M_max_ amplitudes were generally larger during the intervention relative to Baseline, with the largest response observed at Endpoint and a return toward Baseline at Follow-up. The line plot shows MEP motor threshold across timepoints, which was lower throughout the intervention, reaching its lowest value at Endpoint and partially returning toward Baseline at Follow-up. **NOTE**: In all boxplots, data are pooled across tested intensities for MEPs, across target directions for reaching kinematics, or participant-level control cohort comparisons for TRI-HFT. The central line indicates the median, the box represents the interquartile range (IQR), whiskers extend to the minimum and maximum values within 1.5×IQR, and dots represent outliers. Different symbols indicate the mean at each intensity for MEP responses and each target for reaching kinematics. Statistical inference was performed using a bootstrap method (n=10,000 resamples), with asterisks indicating statistical significance.

After four weeks of BCI-ESCS use, the participant exhibited considerable improvements in object manipulation and prehension ability, as assessed by the TRI-HFT (Fig. 4B; Tables S3 and S4) and the Graded Redefined Assessment of Strength, Sensibility, and Prehension (GRASSP) (Fig. 4C; Table S5). Improvements were greater in the BCI-ESCS-targeted right hand, although the left hand, which was not directly targeted by the intervention, showed some functional gains in TRI-HFT (Table S3).

For the BCI-ESCS-targeted right hand, the TRI-HFT 10-object score improved by 9 points at Endpoint and was retained at Follow-up, with notable gains in the ability to manipulate a mobile phone, pop can, paper, and credit card (Fig. 4B). The TRI-HFT rectangular blocks score improved by 17 points at Endpoint and was retained at Follow-up (Fig. 4B). Notably, these right-hand gains after BCI-ESCS use exceeded the average improvements observed in a control cohort of six individuals with chronic SCI who received 40 sessions of conventional therapy over 16 weeks (Fig. 4B; Table S4). The TRI-HFT score improvements exceeded the minimal detectable change (MDC) of 9.1 and 9.2 points for both hands combined in the 10-object and rectangular blocks task, respectively (*37*). These improvements reflect enhanced proximal arm control (shoulder and elbow), wrist coordination and forearm rotation, and grasping and pinching ability (enhanced thumb engagement), collectively supporting more functional motor gains.

Similarly, the GRASSP quantitative prehension scores for both hands revealed improvements in the water bottle, jam jar, and coin tasks (Fig. 4C; Table S5). The 4-point gain at Endpoint surpassed the MDC of 3 points for individuals with SCI (*38*) and was maintained at Follow-up. Beyond the functional outcomes, the ISNCSCI examination revealed neurological-level changes, with a 7-point increase in total motor score and bilateral expansion of the zone of partial preservation in both sensory and motor domains (Table S6).

Improvements in upper-limb coordination were observed during the 3D reaching task (Fig. 4D; Table S7). At Endpoint, both movement smoothness (fewer velocity peaks) and endpoint accuracy (smaller target error) improved compared to Baseline, with only endpoint accuracy retained at Follow-up. Muscle synergy analysis further showed that the number of identified synergies increased from six at Baseline to seven at Endpoint, suggesting increased neuromuscular coordination complexity, before returning to six at Follow-up (Fig. S6). These kinematic and muscle synergy changes suggest that neuromuscular adaptations induced by repeated BCI-ESCS use may have contributed to the functional gains observed in object manipulation and prehension.

Corticospinal excitability was assessed across six timepoints spanning Baseline, the four-week intervention period, and Follow-up (Fig. 4E; Table S7). M_max_-normalized MEP amplitudes in the ECR muscle were generally larger during the intervention than at Baseline, with the largest response observed at Endpoint and a return toward Baseline levels at Follow-up. MEP motor threshold in the ECR was also lower at later longitudinal assessments, reaching the lowest value at Endpoint and partially returning toward Baseline at Follow-up. Complementary changes in MEP amplitudes were observed in the FCR muscle (Fig. S5; Table S7). Although these findings should be interpreted cautiously given the single-participant design, the longitudinal neurophysiological assessments provide preliminary observations consistent with changes in corticospinal excitability associated with BCI-ESCS use.

Longitudinal BCI-ESCS use did not adversely affect other clinical outcomes (“Clinical assessments” section of Supplementary Materials), including the participant’s neurological classification (ISNCSCI, Table S6), spasticity (Modified Ashworth Scale: MAS, Fig. S7), independence (Spinal Cord Independence Measure-Self Care: SCIM-SC, Table S8), quality of life (The International Spinal Cord Injury Quality of Life Basic Data Set: QoL-BDS, Table S9), or pain levels (International Spinal Cord Injury Pain Basic Data Set: ISCIPBDS, Table S10), supporting the feasibility and safety of the intervention.

## DISCUSSION

This first-in-human study provides evidence that an implantable BCI-ESCS system can promote both immediate assistance and sustained functional improvements in upper-limb motor function in an individual with chronic motor-complete cervical SCI. By decoding attempted movement-related ECoG activity to initiate cervical ESCS during functional upper-limb motor tasks, this system realizes a human-in-the-loop control architecture for intention-driven spinal neuromodulation. Three main findings emerged. First, BCI-ESCS provided immediate assistive effects, improving grip force, reaching performance, and object manipulation compared to no stimulation (Fig. 2). Second, BCI-ESCS was associated with increased corticospinal and spinal reflex excitability within the session, whereas tonic stimulation was not (Fig. 3), suggesting the importance of volitional engagement and temporally coordinated stimulation for enhancing descending drive modulation in neuroprosthetic control (*6*, *20*, *21*, *28*, *29*). Third, repeated BCI-ESCS use over four weeks led to meaningful functional improvements exceeding those observed in a reference cohort receiving conventional therapy and established clinical thresholds, together with preliminary evidence of longitudinal changes in corticospinal excitability (Fig. 4). These proof-of-concept findings suggest that leveraging volitional intention to control spinal circuit modulation may represent a promising direction for both neuroprosthetic assistance and motor restoration following paralysis.

Chronic complete SCI is characterized by profound disruption of corticospinal transmission, resulting in the absence of voluntary movement below the injury level (*39*), a state often considered at a spontaneous recovery plateau (*3*, *4*). Our findings provide initial evidence of functional recovery in an individual with chronic tetraplegia. Despite the participant presenting with a motor-complete C4–C5 injury with no detectable voluntary grip force and limited reaching capacity at baseline (Fig. 2; Fig. S1), both BCI-ESCS and tonic stimulation immediately improved grip force, reaching performance, and object manipulation. At the neuromuscular level during reaching, both stimulation modes shifted coordination toward more functionally distinct muscle groupings (Fig. S4) (*32*, *33*). Notably, improvements in grasping and reaching were accomplished through different stimulation contacts, demonstrating the capacity of ESCS to selectively recruit distinct upper-limb muscle groups for task-specific movements (Fig. S2), extending previous reports of assistive effects with tonic spinal stimulation (*7*, *8*, *12–14*). Together, these findings suggest that residual corticospinal drive persists even in chronic motor-complete cervical SCI, which is further supported by the presence of spasticity (Fig. S7), a clinical indicator associated with potential for motor recovery (*40*). ESCS may engage the residual corticospinal connections by recruiting primary afferents in the dorsal roots (Fig. S2), which transsynaptically activate motoneurons and interneurons within the spinal cord (*5*, *18*, *41*, *42*), bringing their membrane potential closer to the firing threshold (*5*, *7*, *43*, *44*). As a result, these neurons become more responsive to residual descending cortical input, effectively lowering the activation threshold for volitional control.

Although both BCI-ESCS and tonic stimulation augmented motor output, they produced different immediate effect profiles. Specifically, BCI-ESCS produced greater grip force and reaching accuracy, whereas tonic ESCS yielded smoother reaching trajectories (Fig. 2). The larger initial direction angle under BCI-ESCS likely reflects an adaptation in movement initiation associated with the abrupt onset of intention-triggered stimulation, consistent with movement overshoots previously reported in BCI-driven stimulation systems (*15*). These assistive effects were achieved using a simple ERD-based trigger contingent upon volitional intention, with an approximately 84% decoding success rate, demonstrating the clinical feasibility of BCI-ESCS and suggesting the potential for further performance gains with more advanced decoding strategies. Taken together, these findings highlight the motor output profile of intention-triggered spinal neuromodulation. By integrating stimulation to motor intention, consistent with previous closed-loop neuroprosthetic strategies (*15*, *19–21*, *45*, *46*), BCI-ESCS may strengthen functional coupling between cortical activity and spinal circuits, supporting facilitation of residual descending pathways for voluntary motor control after SCI (*47–50*).

Beyond providing immediate assistance, BCI-ESCS was associated with changes in corticospinal and spinal excitability following single sessions (Fig. 3B-C), whereas changes in corticospinal excitability were also observed during four weeks of BCI-ESCS use (Fig. 4E). Although these results should be considered in the context of a single participant, they provide preliminary neurophysiological evidence that intention-driven neuromodulation may engage residual sensorimotor pathways. A similar pattern of neurophysiological changes was not observed across tonic ESCS sessions (Fig. 3B-C), although long-term tonic ESCS use has been shown to improve motor function (*12–14*), consistent with the possibility that tonic and intention-triggered stimulation engage activity-dependent plasticity through different mechanisms. The neurophysiological changes observed during BCI-ESCS use are consistent with prior evidence suggesting that intention-driven neuromodulation can promote neuroplasticity and that temporal coupling between descending activity and stimulation may be important (*28*, *29*, *51*). BCI neuromodulation may operate through a Hebbian-like framework, in which repeated temporally coordinated cortical and spinal activation reinforces synaptic efficacy across spared corticospinal connections (*52*, *53*). This principle is also consistent with spike-timing-dependent plasticity (STDP), where the relative timing of cortical and spinal activation drives synaptic potentiation (*43*, *54*). Although BCI-ESCS does not achieve the millisecond-level timing precision characteristic of STDP, repeatedly pairing endogenous cortical activity with subsequent excitation of spinal interneurons may elicit long-term potentiation (LTP)-like increase in corticospinal excitability (*46*). Over time, this repeated pairing could strengthen synaptic efficacy, increase excitatory drive within spinal circuits, and facilitate recruitment of residual descending pathways (*53*).

The functional improvements observed following four weeks of BCI-ESCS use (Fig. 4) likely reflect the cumulative effects of repeated intention-triggered stimulation. Motor performance in object manipulation and reaching, which were immediately enhanced during stimulation (Fig. 2), also showed sustained improvements following four weeks of BCI-ESCS use even in the absence of stimulation (Fig. 4B–D). Notably, the gains in TRI-HFT after BCI-ESCS use surpassed minimal detectable change thresholds (*37*) and even the improvements of a control cohort that underwent 40 sessions of conventional activity-based therapy (Fig. 4B). These functional changes were accompanied by modulation of MEP responses toward the end of the intervention (Fig. 4E), potentially reflecting cumulative build-up observed after a single session (Fig. 3B). The findings are consistent with neuroplastic adaptations (*55*), although the underlying mechanisms remain unknown. At one month follow-up, functional gains persisted (Fig. 4B–D) despite MEP measures returning to baseline (Fig. 4E), suggesting that the retained improvements were not solely associated with changes in corticospinal excitability (*47*). Performance improvements likely also reflect contributions from residual pathways such as the reticulospinal tract (*49*), spinal circuit excitability (*8*), and reorganization of propriospinal networks (*56*).

This proof-of-concept investigation provides converging early evidence across assistive (Fig. 2), short-term (Fig. 3), and longitudinal (Fig. 4) measures, suggesting that BCI-ESCS has a potential to provide neuroprosthetic assistance while also supporting long-term functional improvement. Our findings extend the work of Lorach et al. (*19*), which demonstrated the feasibility of brain-controlled ESCS targeting the lumbosacral spine for lower-limb function, by applying an intention-driven neuromodulation approach at the cervical level for upper-limb function. Together, these studies provide a foundation for broader evaluation of BCI-ESCS and its potential integration with complementary strategies, including pharmacological agents (*57*), cell-based therapies (*58*), or robotic-assisted rehabilitation (*59*).

Several limitations should be acknowledged. First, this proof-of-concept study was conducted in a single individual with chronic motor-complete cervical SCI and therefore cannot establish generalizability. Second, only BCI-ESCS was evaluated longitudinally, preventing direct comparison of long-term effects between BCI-driven and tonic stimulation, whereas comparisons with conventional therapy relied on a historical reference cohort and therefore provide limited evidence regarding comparative benefit. Third, neurophysiological findings were obtained from a limited number of assessments in a single participant and should be interpreted as preliminary observations consistent with, rather than as definitive evidence of, neuroplastic change. Although participant positioning and EMG electrode placement were standardized across assessments and M_max_ normalization was used where available to reduce the influence of peripheral and recording-related variability, day- to-day and state-dependent variability may still have influenced the neurophysiological measurements which should be considered when interpreting the findings.

## MATERIALS AND METHODS

### Trial and participant information

The study was approved by the University of Miami Institutional Review Board (IRB; protocol no. 20190536) and conducted in accordance with the Declaration of Helsinki. The experimental protocol for this study was registered on ClinicalTrials.gov (NCT06533969).

#### Participant information

One individual participated in this study after providing written informed consent. The participant was a male in his 20’s with a history of traumatic C4–C5 SCI (AIS A) who had undergone anterior cervical discectomy and fusion (C3–C5) in 2015. In 2018, the participant underwent implantation of two four-contact ECoG electrodes (Resume II Leads, Medtronic) over the sensorimotor cortex as part of a separate clinical trial evaluating BCI utility in tetraplegia (*23*, *24*). The electrodes were connected to an implantable pulse generator (IPG; Activa PC+S, Medtronic), which enabled real-time recording of cortical signals used by the BCI system (see the “BCI system” section).

The participant was recruited based on the following inclusion criteria: medically stable adult (>18 years old) with chronic (>6 months) traumatic SCI at the C4–C6 level and some residual motor function. Exclusion criteria for the study included contraindications to spinal cord stimulation (for example, wounds near the stimulation site, muscle denervation, unmanaged autonomic dysreflexia, recent botulinum toxin injections), contraindications to cortical stimulation (for example, history of epilepsy/seizures), or surgical ineligibility for ESCS (for example, unfavorable spinal anatomy, anticoagulation therapy, neurological conditions unrelated to SCI, pregnancy).

#### Study design and data reported

This single-case pilot study aimed to assess the therapeutic potential of BCI-ESCS therapy for enhancing upper-limb motor function in chronic SCI. A total of 19 therapy sessions were completed over four weeks (28 days) following ESCS lead implantation (see “Surgical procedure” section of the Supplementary Materials). Each therapy session lasted approximately 30-40 min (40.8±19.3 min) and included at least three motor tasks, each performed for 10 to 30 min. Over the four-week period, the participant completed a total of 16 BCI-ESCS sessions and 3 Tonic-ESCS sessions, enabling comparison of stimulation modes, with a major focus on longitudinal therapeutic effects. The study outcomes focused on evaluating assistive effects (stimulation on/off), short-term effects (pre-post single session), and longitudinal therapeutic effects (recovery of voluntary function).

Assistive effects were assessed by evaluating motor function under three different stimulation conditions: no-stimulation, Tonic-ESCS, and BCI-ESCS. The conditions were tested in a pseudorandomized order to minimize potential order effects. Assessments included maximum grip force measured during MVC (two trials per condition; see “Maximum grip assessment” section of Supplementary Materials), selected TRI-HFT subtests involving manipulation of a mug, mobile phone, dice, and rectangular blocks, as well as performance in a 3D reaching task (see “3D reaching assessment” section of Supplementary Materials).

The single-session effects were assessed in a counterbalanced crossover design to mitigate potential order effects, with two sessions for each of the BCI-ESCS and Tonic-ESCS conditions (Fig. S3). Training trials and tasks were matched across conditions, with sessions lasting 30 min. Neurophysiological assessments, including corticospinal excitability measured through motor cortex stimulation-elicited MEPs and spinal excitability measured through PRM reflexes elicited by ESCS, were conducted before and after each therapy session. Additionally, functional motor performance was evaluated using the 9HPT and the BBT (see “Clinical assessments” section of Supplementary Materials) due to their sensitivity to subtle motor changes over short timeframes.

To assess longitudinal effects, functional and clinical evaluations were conducted at three time points: Baseline, before the longitudinal BCI-ESCS therapy period; Endpoint, at the end of the therapy period; and Follow-up, one month after lead explantation (Fig. S3). Corticospinal excitability was assessed at six timepoints: Baseline (2 days before ESCS lead implantation), Day 6, Day 10, Day 21, Endpoint (Day 22), and Follow-up (4 weeks after ESCS lead explantation) (Fig. S3). The primary outcome measure was functional improvement assessed using the TRI-HFT. Secondary outcomes included changes in GRASSP scores, neurophysiological measures of corticospinal excitability, kinematics and muscle synergies during a 3D reaching task (see “3D reaching assessment” section of Supplementary Materials), and clinical assessments of pain, quality of life, independence, spasticity, and ISNCSCI (see “Clinical assessments” section of the Supplementary Materials).

### Epidural spinal cord stimulation (ESCS) system

ESCS was administered using a constant-current electrical stimulator (DS8R, Digitimer), a multiplexer (D188, Digitimer), and a custom-built control unit to regulate the pulse timing, amplitude, and output channel selection (*13*) (Fig. S8). To interface the stimulator with the percutaneous leads, the system was connected to an accessory kit (355531, Medtronic) via a Medusa electrode cable (Threshold NeuroDiagnostics). Selected contacts on the percutaneous leads served as cathode electrodes, and a 2x4" anode electrode (Natus) was placed over the left (contralateral) iliac crest. Each stimulation pulse was delivered as a symmetrical biphasic waveform with a 200 µs pulse width and 10 µs interphase interval (Fig. 5).

**Fig. 5.**
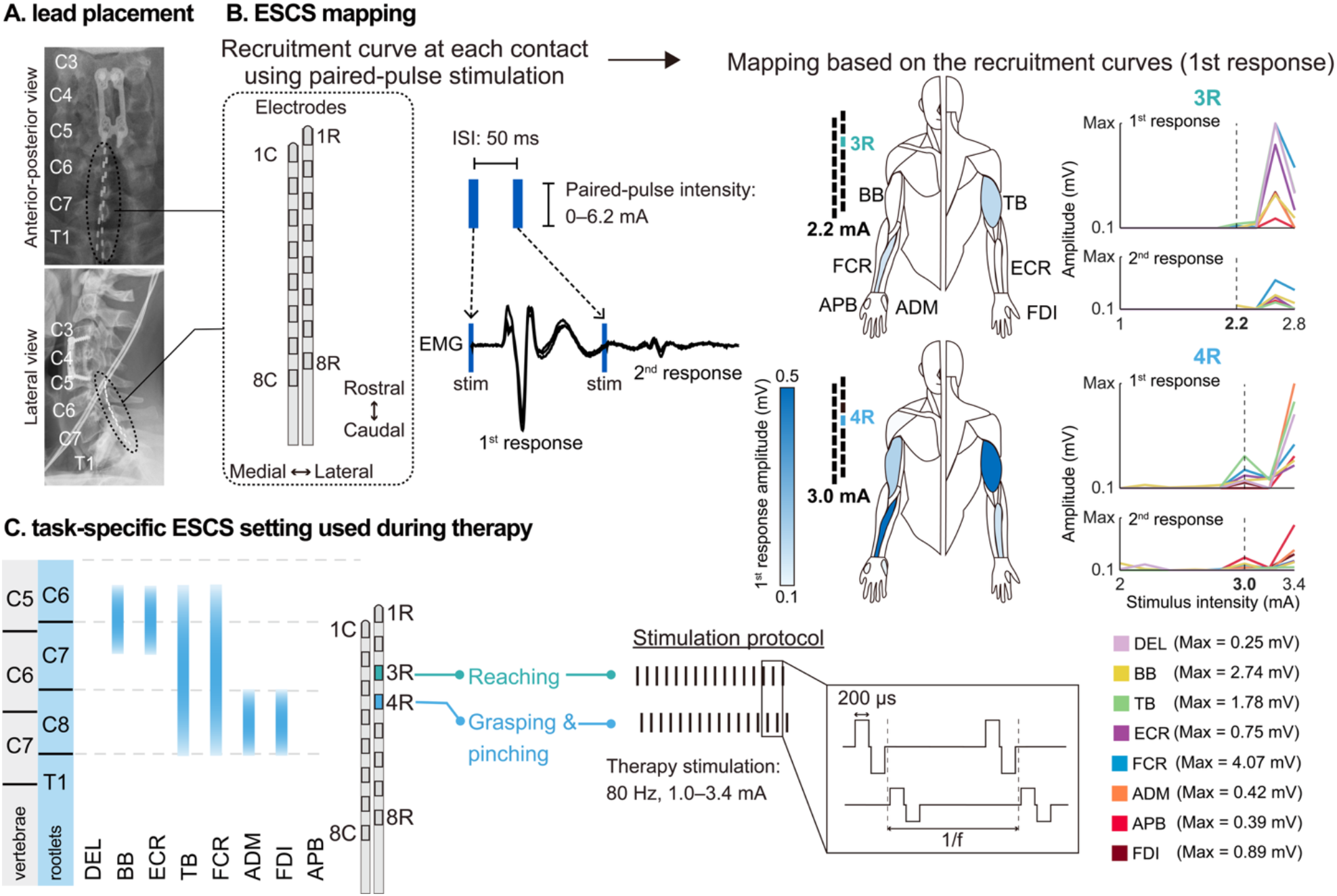
ESCS lead placement and spinal mapping for muscle recruitment. **(A)** Lead placement: Post-operative X-ray images showing the positioning of the percutaneous epidural spinal cord stimulation (ESCS) leads in the cervical epidural space. The leads were implanted over the dorsal roots at C6–T1 to selectively target upper-limb muscles. **(B)** ESCS mapping: Recruitment curves were obtained by applying paired-pulse stimulation (interstimulus interval (ISI): 50 ms) at increasing intensities (0–6.2 mA) to each contact. EMG recordings were used to assess evoked muscle responses. Recruitment curves for contacts 3R and 4R illustrate muscle-specific activation patterns, with the second response showing post-activation depression, confirming activation via primary afferent fibers. **(C)** Task-specific ESCS settings used during therapy: Stimulation parameters were optimized to selectively modulate reaching, grasping, and pinching movements, with stimulation intensities ranging from 1.0 to 3.4 mA at 80 Hz and a biphasic pulse width of 200 µs. These settings ensured targeted muscle activation while minimizing unintended co-contraction.

#### Segment-level muscle recruitment

Using intraoperative fluoroscopy as an initial anatomical reference (Fig. S9), we conducted a spinal mapping session within the first few days after implantation to determine segment-level muscle recruitment profiles. Muscle recruitment curves were generated by applying paired-pulse stimulation with a 50-ms interstimulus interval (*30*, *31*). For each contact, the stimulation intensity was gradually increased from 0 mA until evoked responses were observed in all right-hand muscles or any contralateral left-sided responses appeared. Stimulation intensity did not exceed 6.2 mA. From the recruitment curves, we identified contacts and stimulation intensities that produced selective muscle recruitment through activation of primary afferent fibers (Fig. 5), as indicated by post-activation depression of the second responses (*30*, *31*). These identified contacts and intensities served as references for subsequent therapy sessions, with brief spinal mapping assessments conducted before each session to verify stable recruitment. Given that deviations in recruitment patterns were observed during pre-therapy verification on day 7 post-implant, we repeated complete spinal mapping and acquired additional cervical plain radiography (Fig. S9) to verify lead position and re-optimize stimulation parameters. Consequently, we updated optimal contacts for functional tasks such as reaching (shifted from 4R to 3R) and grasping (shifted from 5R to 4R) (Fig. S3), corresponding to the lead displacement (Fig. S9). Daily spinal mapping verification in subsequent days confirmed stable recruitment profiles throughout the remainder of therapy, which aligns with prior evidence showing minimal lead displacement after the second week (*13*).

#### ESCS optimization for intervention sessions

For each therapy task, ESCS contacts were selected based on segment-level muscle recruitment mapping, with stimulation parameters (frequency and intensity) optimized to maximize support during volitional movements while preventing involuntary muscle twitching. Specifically, stimulation contacts that preferentially recruited upper-arm muscles were selected for reaching tasks and contacts that preferentially recruited forearm and hand muscles were selected for grasping and pinching tasks (Fig. 5). At the beginning of each therapy session, stimulation contacts were adjusted to ensure optimal neuromodulation, and alternative contacts were used if necessary to enhance motor performance (Fig. S3). For each selected contact, stimulation frequencies ranging from 40 to 100 Hz were tested (*13*), with 80 Hz identified as optimal for maximizing assistive benefits. Stimulation intensities were set using a functional, task-based criterion, and titrated to facilitate volitional movement while remaining below the level that produced overt movement at rest (*8*, *13*). The intensities were marginally adjusted throughout therapy, ranging from 1.0 to 3.4 mA.

### BCI system

#### ECoG acquisition system

Cortical activity was recorded using two four-contact electrodes (Resume II Leads, Medtronic) implanted over the hand knob region of the sensorimotor cortex during previous clinical trials (*23*, *24*) (Fig. 6). These electrodes were subcutaneously connected to an IPG (Activa PC+S, Medtronic) that enables the recording of two bipolar time-series channels (Ch 0-3 and Ch 8-10) (Fig. S8). In this study, the IPG was used for ECoG signal acquisition to support real-time BCI decoding. Single-pulse motor cortex stimulation was also delivered via the IPG for MEP assessment, as described in the “Corticospinal excitability” section. The acquisition system applied a 0.5-100 Hz band-pass pre-amplifier filter and sampled data at 200 Hz, with an internal buffer of 400 ms (*60*).

**Fig. 6.**
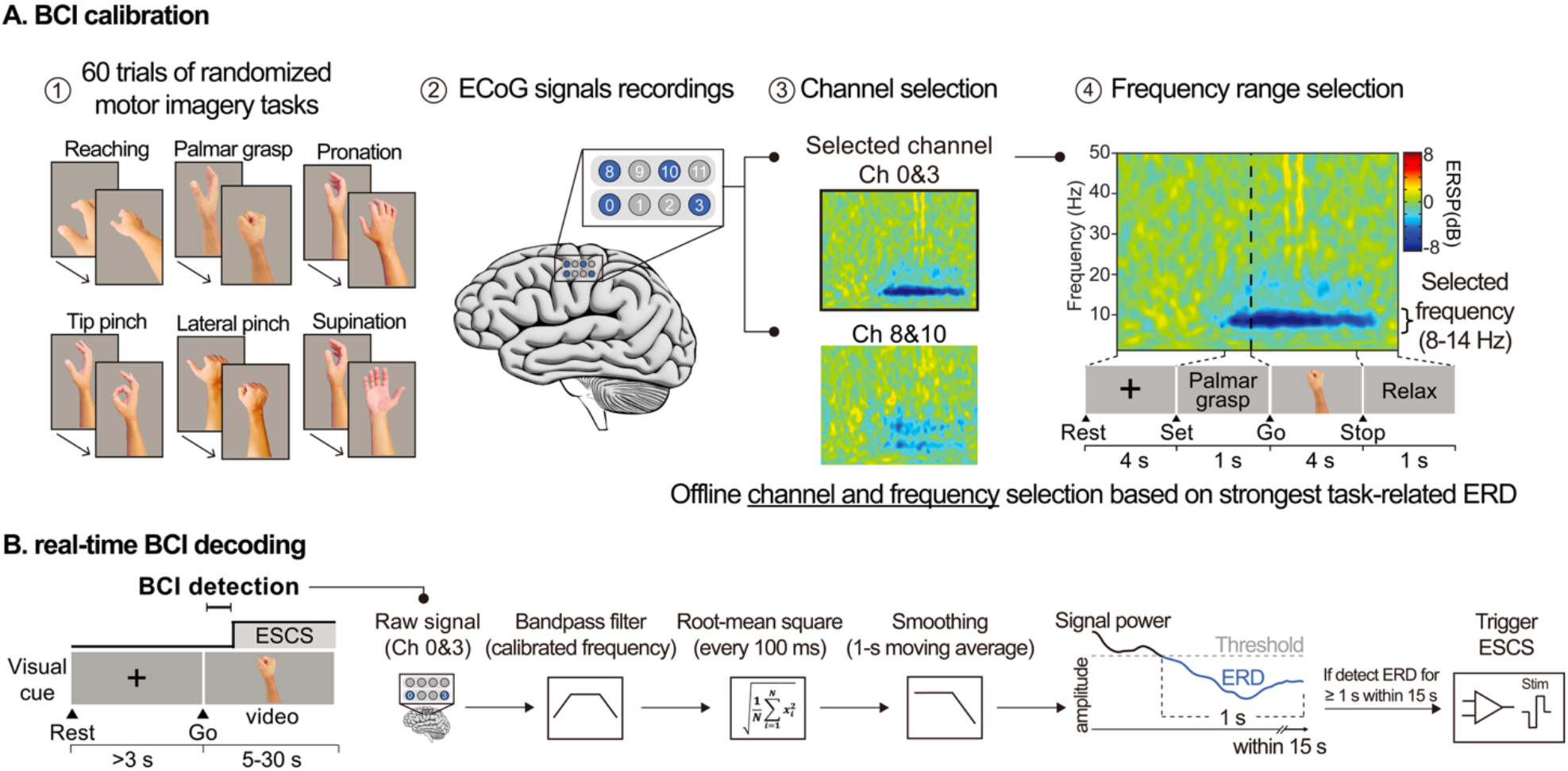
BCI calibration and real-time decoding for BCI-ESCS therapy. **(A)** BCI calibration process. The participant performed 60 trials of randomized motor imagery tasks, including reaching, palmar grasp, pronation, tip pinch, lateral pinch, and supination. ECoG signals were recorded from implanted electrodes, and offline analysis was conducted to determine the optimal ECoG channel and frequency range based on event-related desynchronization (ERD). Channel selection was performed by comparing activation patterns across electrodes, and the frequency range (8-14 Hz) was selected based on the strongest task-related ERD. **(B)** Real-time BCI decoding during therapy. Raw ECoG signals from the selected channels (Ch 0 and Ch 3) were processed in real-time using a band-pass filter (calibrated frequency) and converted into a root-mean-square (RMS) value every 100 ms. A 1-second moving average smoothing was applied to track signal power. If the ECoG power remained below a predefined ERD threshold for at least one second within a 15-second window, the BCI system triggered ESCS to support the intended movement.

#### BCI decoding

The BCI system was adapted from previous studies (*25–29*) to detect motor imagery of upper-limb movements (Fig. 6) by analyzing real-time changes in ECoG signal power. This signal was then used to trigger task-specific ESCS during activity-based therapy (see the “BCI-ESCS therapy protocol” section).

Before therapy (baseline), a BCI calibration session was conducted to determine the optimal channel and frequency. During this session, the participant performed 10 motor imagery trials for each of the following movements: reaching, grasping, supination, pronation, lateral pinching, and pulp pinching. From these 60 total trials, time-frequency spectrograms were computed to identify the optimal ECoG channel (Ch 0-3) and frequency range (8-14 Hz) exhibiting the strongest event-related desynchronization (ERD) during motor imagery tasks (*26–29*) (Fig. 6). These calibrated features were subsequently used for BCI decoding across all motor imagery tasks.

To decode motor imagery during BCI-ESCS therapy, a one-degree-of-freedom classifier was developed to detect the transition between rest and active motor imagery states (*25–29*). A single decoder was used across all motor imagery tasks, such that stimulation could be activated by imagining any of the trained movements. Decoding was based on ERD activity from the calibrated channel (Ch 0 and Ch 3) and frequency band (8-14 Hz) identified during the baseline BCI calibration session (Fig. 6A). For each trial, video cues (Fig. 6B) instructed the participant on the specific task to guide task execution during therapy. Specifically, the BCI system acquired ECoG signals from the selected channel and applied a band-pass filter within the calibrated frequency range. The root-mean-square (RMS) value of the filtered signal was computed every 100 ms and subsequently smoothed using a 1-second moving average window to obtain the signal power. To trigger ESCS, the BCI system monitored the ECoG signal power within a 15-second window after the participant was instructed to imagine the corresponding task (Go cue, Fig. 6). If the signal power remained below a predefined threshold for at least one second (*25–29*), the BCI system triggered ESCS for a task-specific, pre-programmed duration (4–25 s) to assist the participant in performing the corresponding activity-based therapy task. At the end of this stimulation window, ESCS automatically returned to the off state and remained off until the next trial. The threshold was determined prior to each therapy session during a practice period. The experimenter initially adjusted the signal power threshold to distinguish rest from motor imagery states, and the threshold was readjusted periodically throughout sessions as needed to compensate for drift (*25–29*). Conversely, if motor imagery was not detected, ESCS was not triggered, and the participant remained at rest before proceeding to the next task. BCI performance was quantified as the success rate, calculated as the proportion of detected motor imagery (successful trials) out of the total number of trials, resulting in a mean success rate of 83.8±4.2%. This success rate remained stable across therapy sessions (Spearman rank correlation between session number and success rate: *ρ*≈0*, p*>0.999, df=14), with no significant differences found across motor tasks (Kruskal-Wallis test: H(5)=0.757, *p*=0.980).

### BCI-ESCS task protocol

Therapy was conducted by a physical therapist and two BCI-ESCS operators, focusing on restoring right upper-limb function through activity-based training with BCI-triggered ESCS. The physical therapist instructed the participant through a series of motor tasks with assistance provided as needed, while the BCI system continuously monitored motor imagery and triggered ESCS in real time to facilitate volitional movement.

At the start of each task, a visual cue corresponding to the intended movement (reaching, grasping, pronation, supination, lateral pinching, and pulp pinching) was presented on a monitor (Fig. 7A). Upon receiving the “Go” cue, the participant attempted to imagine the instructed movement. When motor imagery was detected (see the “BCI decoding” section), the BCI system triggered ESCS, providing stimulation to enhance movement execution. If necessary, the therapist provided minimal assistance to the upper limb, thereby enabling the participant to move in the intended direction with voluntary effort and less compensation.

**Fig. 7.**
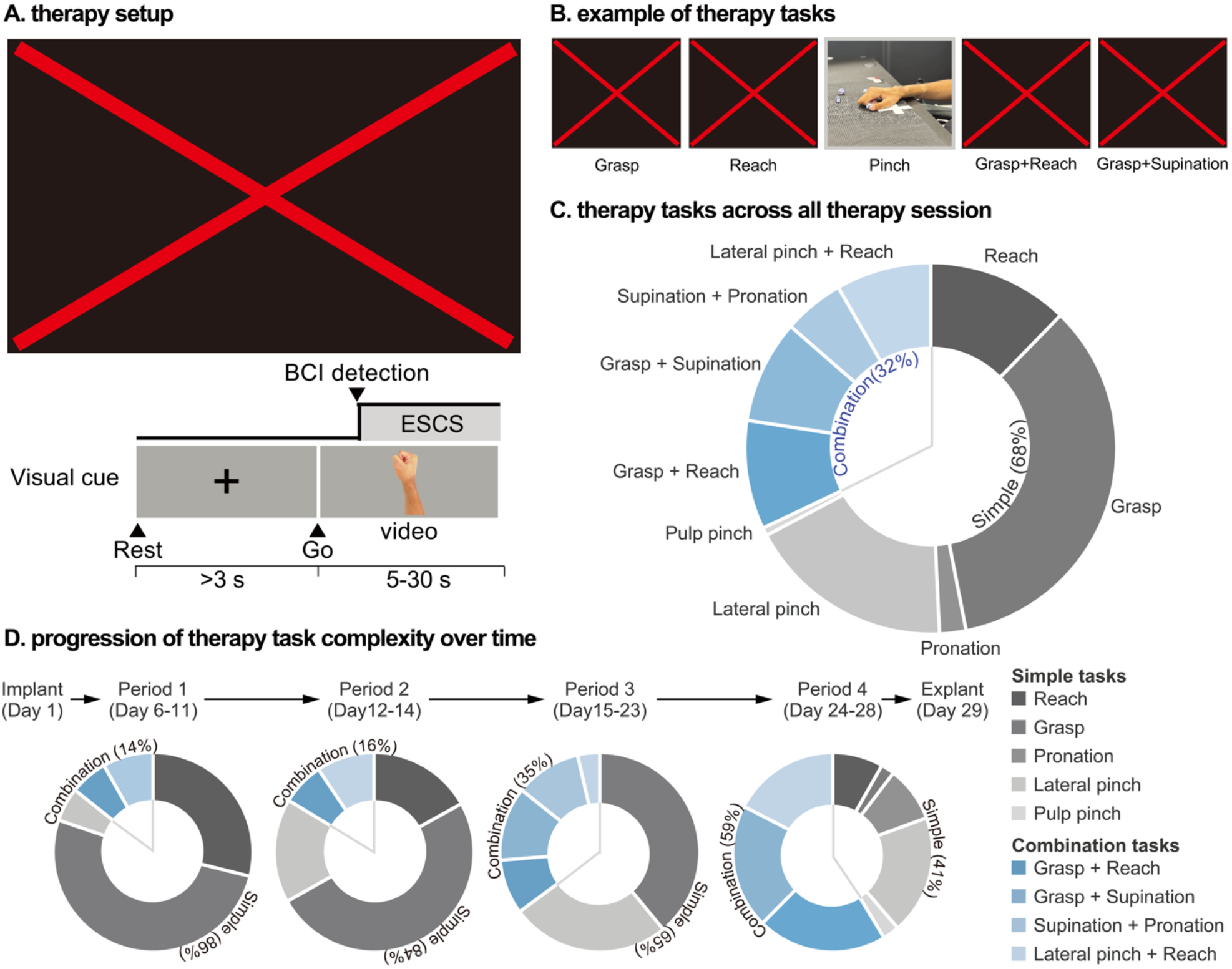
BCI-ESCS setup and task progression over the long-term intervention. **(A)** Therapy setup: The participant sat in a wheelchair facing a monitor displaying visual cues corresponding to the instructed motor tasks. Upon receiving the “Go” cue, the participant attempted to imagine the instructed movement while the BCI system detected motor intention and triggered ESCS in real time. The stimulation remained active throughout the task execution phase. **(B)** Examples of therapy tasks. The participant engaged in functional hand and arm movements, including grasping, reaching, pinching, and combined movements such as grasp + reach and grasp + supination, mimicking real-life functional tasks. **(C)** Task distribution across all therapy sessions. Throughout the four weeks (28-day) therapy period, the participant performed a variety of motor tasks, including simple isolated movements (reaching, grasping, pronation, supination, lateral pinching, pulp pinching) and complex combination movements (grasp + reach, grasp + supination, supination + pronation, and lateral pinch + reach). Simple movements accounted for 68% of total tasks, whereas combination movements comprised 32%, reflecting a progressive increase in task complexity aimed at improving functional motor coordination. **(D)** Progression of therapy task complexity over time. The proportion of combination tasks increased throughout the four-week therapy period. Initially, simple tasks dominated the sessions (86% early on), but as therapy progressed, the participant performed a greater proportion of complex combination tasks, reaching 59% by the final sessions.

To progressively enhance motor function, task difficulty was adjusted throughout the intervention. Initially, the participant engaged in simple isolated movements (for example, reaching or grasping) (Fig. 7B-C). As performance improved, more complex combination tasks (for example, reaching and grasping simultaneously) were introduced (Fig. 7D). Training was further enriched using objects of varying weight, size, and texture (for example, cups, blocks, balls) to enhance sensorimotor engagement. Additionally, task parameters such as movement height and distance were modified to challenge motor control. In later stages of therapy, the participant practiced functional movements relevant to daily living, such as mimicking drinking or eating, to promote practical skill acquisition.

### Tonic-ESCS as a control for assistive and short-term effects

Tonic-ESCS was implemented as a control condition to evaluate the assistive and short-term (single session) effects of BCI-ESCS use. During the Tonic-ESCS condition, stimulation was delivered continuously (without BCI triggering) throughout the session. Stimulation parameters were optimized before each task, following the same procedure used in the BCI-ESCS condition, ensuring that stimulation facilitated movement without excessive involuntary muscle activation.

To minimize variability in the short-term assessments, the BCI-ESCS and Tonic-ESCS conditions included matched therapy tasks and durations under the guidance of a physical therapist.

### Hand function assessments

The TRI-HFT and GRASSP assessments were administered by a licensed physical therapist who also served as the training therapist in this study. To mitigate potential rater bias, an independent rater, trained in TRI-HFT and GRASSP but not involved in the intervention, scored performance from video recordings. Any discrepancies between the two raters were resolved through discussion to reach consensus.

#### Toronto Rehabilitation Institute-Hand Function Test (TRI-HFT)

The TRI-HFT is a standardized assessment of unilateral gross motor function of the upper extremity, with an emphasis on pinch and palmar grasp. We assessed two components: (1) 10-object manipulation, where individuals are evaluated on their ability to manipulate ten everyday objects, and (2) rectangular blocks, where individuals are evaluated on their ability to manipulate blocks with varying weights and friction levels (*61*). We used a 3D-printed version of the TRI-HFT, which has been validated in individuals with SCI (*62*). Each object is scored on a scale from 0 to 7, with higher scores reflecting better function.

To assess the effects of our four-week BCI-ESCS intervention, we compared TRI-HFT outcomes to control data from six individuals with chronic SCI (n=2: C5–C6 complete SCI and n=4: C4–C7 incomplete SCI; 5.7±3.1 years since injury) who underwent conventional activity-based therapy without neuromodulation at the Rehabilitation Engineering Laboratory (KITE Research Institute - University Health Network). The training protocol and partial dataset are described by Kapadia et al. (*63*), where participants completed 40 one-hour therapy sessions over 16 weeks. In contrast, the individual in our current study completed 19 sessions of approximately 40 min of BCI-ESCS use combined with comparable activity-based rehabilitation. The control comparison evaluated whether the TRI-HFT improvements in our participant with a motor-complete SCI exceeded those observed in the control group.

#### Graded Redefined Assessment of Strength, Sensibility and Prehension (GRASSP) Version 1.0

The GRASSP v.1.0 is a comprehensive and sensitive measure of the upper-limb impairment and function in the cervical SCI population, which was used to assess three domains of recovery: sensation, strength, and prehension (*64*, *65*).

### Neurophysiological assessments: corticospinal and spinal excitability

All neurophysiological assessments were performed with the participant seated in a wheelchair using the same seat configuration and upper-limb positioning across conditions and sessions. The seat angle was maintained at approximately 15°, the seat-to-backrest angle at 90°, elbows at approximately 90° flexion supported by the armrests, and palms facing downward. Background EMG was monitored to ensure the absence of overt muscle activity throughout the assessments. The RMS amplitude of background EMG activity was also quantified over the 50-ms period preceding each MCS or ESCS stimulus during corticospinal and spinal excitability assessments, respectively. Background EMG values (Table S11) were below 5 µV across assessments (*66*) and were considerably lower than MEP and PRM reflex response amplitudes (Fig. 3B-C; Fig. 4E), supporting that responses were obtained under resting conditions.

### Corticospinal excitability

We measured corticospinal excitability using MEPs of the ECR (Fig. 3B; Fig. 4E) and FCR (Fig. S5) muscles elicited by single-pulse MCS delivered through the subdural ECoG electrode. An IPG (Activa PC+S, Medtronic) delivered 450-µs pulses in bipolar configuration, with contact 9 as the anode and contact 10 as the cathode. This contact configuration was selected based on our preliminary investigation, as it elicited MEPs with peak-to-peak amplitudes above 50 µV (*67*) in the ECR muscle at the lowest stimulation intensity. Peak-to-peak MEP amplitudes were recorded across stimulation intensities from 4.0 to 10.5 V, with five stimuli per intensity, and amplitudes were pooled across intensities and sessions for analysis. MEP motor threshold in the ECR muscle was also assessed, with a lower value indicating greater corticospinal excitability (*68*). Motor threshold was defined as the lowest stimulation intensity that elicited a peak-to-peak MEP response exceeding 50 µV in at least half of the trials (*67*). Single-session effects were assessed immediately before (Pre) and after (Post) each BCI-ESCS or Tonic-ESCS session using a counterbalanced crossover design (Fig. S3). Pre-intervention MEP amplitudes between BCI-ESCS and Tonic-ESCS conditions did not reach statistical significance in either muscle (ECR: *p*=0.590, d=-0.254; FCR: *p*=0.056, d=-0.433; Table S2). For longitudinal assessments, MEPs were assessed across six timepoints: Baseline (2 days before implantation), Day 6, Day 10, Day 21, Endpoint (Day 22), and Follow-up (4 weeks after explantation) (Fig. S3). The MEP recordings at Day 6, Day 10, Day 21, and Day 22 also served as the Pre measurements for the corresponding single-session BCI-ESCS or Tonic-ESCS assessments.

### Spinal excitability

Spinal excitability was assessed before and after each BCI-ESCS and Tonic-ESCS session. Posterior root muscle (PRM) reflexes of the ECR (Fig. 3C) and FCR (Fig. S5) muscles were elicited by paired-pulse ESCS using the 4R contact, which was selected as it simultaneously induced responses in both muscles at the lowest stimulation intensity. Post-activation depression observed in paired-pulse responses (Fig. S2) confirmed activation via sensory fibers, which transsynaptically activate motoneurons to produce PRM reflexes (*30*, *31*). Peak-to-peak PRM reflex amplitudes were recorded at gradually increasing intensities, with three paired-pulse stimuli per intensity. Because paired-pulse ESCS intensities differed between sessions due to minor ESCS lead displacement (Fig. S3; Fig. S9), the intensities for PRM analyses were referenced to a session-specific Pre-measurement PRM reflex motor threshold (MT), defined as the lowest intensity at which the first response exceeded 50 µV peak-to-peak (*69–71*) in at least half of the trials. To capture a comparable physiological range spanning subthreshold levels, activation threshold, and the rising slope of the recruitment curve, responses from MT-0.2 mA to MT+0.6 mA were pooled across the two sessions per condition for analysis. MT values for the first session were BCI: 2.8 mA and Tonic: 2.2 mA, and for the second session were BCI: 2.4 mA and Tonic: 2.2 mA. In addition, changes in PRM reflex motor threshold from before (Pre) to after (Post) each single-session intervention were assessed in the ECR muscle as a measure of spinal excitability.

#### Maximum motor responses

Maximum M-wave (M_max_) was assessed in the ECR muscle for normalization of the MEP and PRM reflex amplitudes to reduce the influence of peripheral excitability and recording-related variability across assessments (*34*). M_max_ was obtained by supramaximal electrical stimulation of the radial nerve using a constant-current electrical stimulator (DS7R, Digitimer). Single monophasic rectangular pulses with a 1-ms pulse duration were delivered through surface stimulation electrodes placed over the radial nerve on the lateral aspect of the arm above the elbow. The optimal stimulation site was identified at the beginning of each session and maintained throughout. Stimulation intensity was progressively increased until the M-wave amplitude reached a plateau, defined as no further increase in the response amplitude with increasing intensity (*34*). The stimulation intensity used for M_max_ assessment was then set to 120% of the plateau intensity. Five M_max_ responses were elicited at each measurement timepoint. The mean peak-to-peak M_max_ amplitude at each timepoint was then used to normalize the corresponding MEP and PRM reflex amplitudes.

### Safety

The study was designed to prioritize participant safety, with continuous monitoring by a multidisciplinary team of neurosurgeons, physical therapists, and medical staff. Post-operatively, we conducted daily wound care. A standard post-operative dressing was applied over the cephalocaudal incision, which was sealed with surgical glue, and changed at least five times per week. A silver-infused antimicrobial barrier wound dressing (Silverlon Island Dressing ID-46) was applied over the extensions that exited from the participant’s back and changed approximately every three days (*13*). There was no evidence of infection at the lead exit sites.

All adverse events were recorded, reported to the IRB, and managed by the medical team. There were no serious adverse events reported during the study, and none were related to the BCI-ESCS use. The participant experienced a urinary tract infection during the study that was promptly resolved. We also did not observe any autonomic dysreflexia episodes, and overall vital signs (blood pressure, heart rate, and oxygen saturation) remained stable throughout the study (Fig. S10). A post-session drop in blood pressure was observed during the early therapy sessions (Fig. S10), which was determined by the study team to be most likely due to changes in posture during prolonged upright sitting. Furthermore, improvement in motor function (ISNCSCI in Table S6, TRI-HFT in Table S3, and GRASSP scores in Table S5) suggests the absence of neurological deterioration. Spasticity (Fig. S7) and pain levels (Table S10) also remained stable, with no signs of deterioration or relief, suggesting that the intervention was safe.

### Statistical analysis

Statistical comparisons were performed using a bootstrap resampling method to obtain distributions of mean differences, from which 95% confidence intervals (CIs) were derived (*13*). Statistical significance was determined by rejecting the null hypothesis of no mean difference when the CI excluded zero (p<0.05). Where multiple comparisons were conducted, Bonferroni correction was applied to adjust p-values accordingly. Effect sizes were reported as Cohen’s d.

For assistive effects, grip strength and EMG activity during maximal voluntary contraction were compared between conditions (no-stim, Tonic-ESCS, and BCI-ESCS) using effect sizes as descriptive indicators only, given the limited sample size. Reaching kinematics (velocity peaks, target error, and initial direction angle) were analyzed on data pooled across four reaching directions (left, right, upward, and center), with three pairwise comparisons among conditions.

For single-session neurophysiological effects, MEP and PRM reflex amplitudes were analyzed on data pooled across all tested stimulation intensities and across two sessions within each condition, with six pairwise comparisons among four groups (BCI-ESCS Pre, BCI-ESCS Post, Tonic-ESCS Pre, Tonic-ESCS Post).

For longitudinal effects, MEP amplitudes were analyzed on data pooled across all tested stimulation intensities across six timepoints (Baseline, Day 6, Day 10, Day 21, Endpoint, and Follow-up; Fig. S3), with fifteen pairwise comparisons. Reaching kinematics were analyzed on data pooled across four reaching directions (left, right, upward, and center) across the three timepoints (Baseline, Endpoint, and Follow-up). Additionally, functional outcome changes in TRI-HFT were compared against a control cohort.

## Supplementary Materials

### List of Supplementary Materials

- Supplementary methods: Surgical procedure

- Supplementary methods: Electromyography acquisition system

- Supplementary methods: Clinical assessments

- Supplementary methods: Maximum grip assessment

- Supplementary methods: 3D reaching assessment

- Fig. S1. Baseline neurophysiological evidence supporting motor-complete classification.

- Fig. S2. Spinal mapping for optimizing ESCS contacts.

- Fig. S3. Timeline of stimulation parameters and assessments throughout BCI-ESCS therapy.

- Fig. S4. Assistive effects of BCI-ESCS and Tonic-ESCS on muscle synergies.

- Fig. S5. Neurophysiological responses in the FCR muscle.

- Fig. S6. Longitudinal effects of BCI-ESCS use on muscle synergies.

- Fig. S7. Longitudinal assessment of upper limb spasticity using the Modified Ashworth Scale (MAS).

- Fig. S8. BCI-ESCS system integration.

- Fig. S9. Epidural Spinal Cord Stimulation (ESCS) lead position during early implantation period.

- Fig. S10. Stability of vital signs during the study period.

- Table S1. Statistical results of assistive effects.

- Table S2. Statistical results of single-session effects on corticospinal and spinal excitability.

- Table S3. Longitudinal effects of BCI-ESCS therapy on the Toronto Rehabilitation Institute– Hand Function Test (TRI-HFT) 10-object manipulation and rectangular block scores.

- Table S4. Statistical results of longitudinal effects on the Toronto Rehabilitation Institute–Hand Function Test (TRI-HFT) 10-object manipulation and rectangular block scores.

- Table S5. Longitudinal effects of BCI-ESCS therapy on the Graded Redefined Assessment of Strength, Sensibility and Prehension (GRASSP) scores.

- Table S6. Longitudinal effects of BCI-ESCS therapy on the International Standards for Neurological Classification of Spinal Cord Injury (ISNCSCI) upper extremity motor scores (UEMS) and zones of partial preservation (ZPP) assessments.

- Table S7. Statistical results of longitudinal effects on reaching kinematics and corticospinal excitability.

- Table S8. Longitudinal effects of BCI-ESCS therapy on the Spinal Cord Independence Measure-Self Care (SCIM-SC) scores.

- Table S9. Longitudinal effects of BCI-ESCS therapy on the International Spinal Cord Injury Quality of Life Basic Data Set (QoL-BDS) scores.

- Table S10. Longitudinal effects of BCI-ESCS therapy on the International Spinal Cord Injury Pain Basic Data Set (ISCIPBDS) changes in pain prevalence, number of pain problems, and pain interference scores.

- Table S11. Background EMG during single-session and longitudinal neurophysiological assessments.

## Data Availability

All data produced in the present study are available upon reasonable request to the authors.

## Acknowledgments

We are grateful to Letitia Fisher for her valuable contributions as the clinical coordinator and Nelson Rodriguez for assistance with graphic design. We thank Deena Cilien, DPT and Cristina Segredo, DPT for their expert physical therapy support and for assisting with clinical assessments throughout the study. We also thank Felipe Monteiro, MD and Carlos Blandino for surgical support and Luigi Borda for assistance with the technical design. We extend our sincere gratitude to the research participant for their time, dedication, and insightful contributions throughout the study.

## Author contributions

Conceptualization: AS, RAF, MC, MM

Data curation: AS, RAF

Formal analysis: AS, RAF, AY, ZB, HS, NeM

Funding acquisition: MM, WDD

Investigation: AS, RAF, AY, ZB, HS, RdeF, NiM, NiD, JJ, AP, MEI, JDG, JGC

Methodology: AS, RAF, AY, ZB, HS, RdeF, NiM, EP, MC, JGC, MM

Project administration: RAF, KD, GSP, ST, MM

Resources: NaD, VZ, MRP

Software: RAF, MC

Supervision: MM

Validation: NiD

Visualization: AS, RAF, AY, and ZB

Writing – original draft: AS, RAF, AY, ZB, HS, MM

Writing – review & editing: AS, RAF, MM

## Data, code and materials availability

Data supporting the findings of this study are openly available at https://doi.org/10.5281/zenodo.20752133. To protect participant confidentiality, raw data requests should be made formally to MM, AS or RAF Reasonable requests will be fulfilled and executed through a material transfer agreement between the University of Miami and the requesting party, taking into account any existing sponsored research agreements with the funders. Custom code used to process, analyze, and visualize the data, as well as the ESCS control interface, are available upon reasonable request to MM, AS, or RAF, and will be subject to a suitable code sharing agreement, in compliance with University of Miami policies and any applicable third-party licensing or funding agreements.

