## Supplementary Materials for "Brain-Controlled Epidural Spinal Stimulation for Upper-Limb Motor Function after Tetraplegia"

### Surgical procedure

Prior to surgery, the participant underwent pre-operative screening, including laboratory tests, imaging, prophylactic antibiotics, and a thorough discussion with the research and medical teams over the risks involved. Pre-operative imaging included a non-contrast computed tomography (CT) scan of the head, cervical, and thoracic spine. The ESCS system required temporary surgical placement of two eight-contact percutaneous leads (PN 977A260, Medtronic) in the epidural space of the spinal cord for 28 days, after which the same surgical team removed the leads. During surgery, the participant was placed under general anesthesia, and a head fixation device stabilized the participant's head and neck. A short cephalocaudal incision was made along the upper thoracic spine midline, and an epidural needle was inserted under fluoroscopic guidance into the interlaminar space, leveraging evidence that rostral entry points minimize displacement (*72*). Both leads were placed unilaterally on the right side, with the most rostral tips of the leads placed just caudal to the level of injury, just below the C5 vertebra, with the lateral lead more rostral than the medial lead by one contact (Fig. 1; Fig. S9). Lead positioning was guided by intraoperative fluoroscopy and electrophysiological feedback of muscle action potentials, ensuring selective recruitment of ipsilateral motor pools (13). During this monitoring, the lead was temporarily connected to the intraoperative monitoring system (Natus XLTEK Protektor 32) via 60-cm extensions (PN 3708160, Medtronic), which were removed after testing. The leads were then anchored to the fascia and tunneled subcutaneously to a distant counter incision where the extension leads were externalized for use in later stimulation procedures coupled with neural signal recordings. Percutaneous leads were removed 28 days after implantation under monitored anesthesia care by reopening the cephalocaudal incision and releasing the anchors.

### Electromyography acquisition system

Electromyographic (EMG) activity was recorded during maximum grip assessments (see "Maximum grip assessment" of the Supplementary Materials), the ESCS spinal mapping session (see the "Segment-level muscle recruitment" section), and neurophysiological assessments (see the "Neurophysiological assessments: corticospinal and spinal excitability" section). EMG signals were recorded from the right deltoid (DEL), biceps brachii (BB), triceps brachii (TB), extensor carpi radialis (ECR), flexor carpi radialis (FCR), abductor digiti minimi (ADM), abductor pollicis brevis (APB), and first dorsal interosseous (FDI) muscles. For each muscle, the muscle belly was identified, and the electrode location was marked on the skin. The marked areas were shaved using disposable razor blades and cleaned with alcohol swabs before placing bipolar surface EMG electrodes (Ag/AgCl, H124SG, Kendall) over each muscle belly. An approximate interelectrode distance of 20 mm was used in accordance with Surface EMG for Non-Invasive Assessment of Muscles (SENIAM) recommendations (*73*). Ground electrodes were placed over the ulnar styloid process and the lateral epicondyle. All signals were amplified (×1,000), band-pass filtered (15–1,000 Hz) using bio-amplifiers (D360R, Digitimer), and digitized (4,000 Hz) with an analog-to-digital converter (Power 1401, Cambridge Electronic Design).

### Clinical assessments

***The International Standards for Neurological Classification of Spinal Cord Injury (ISNCSCI).*** The ISNCSCI is a standardized classification tool for SCI used to systematically assess sensory and motor levels of injury to determine the neurological level of the injury and severity (completeness) of the SCI (*74*). ISNCSCI classification at Baseline and Endpoint was performed by a dedicated assessor specializing in standardized neurological evaluations, ensuring consistency and minimizing inter-rater variability. Upper extremity motor scores (UEMS) and sensory and motor zones of partial preservation (ZPP) were reported.

***The International Spinal Cord Injury Pain Basic Data Set (ISCIPBDS) version 2.0.*** The ISCIPBDS version 2.0 was developed to collect a minimal amount of standardized information necessary for determining interference of pain with physical and emotional function and sleep, probable pain diagnosis, location, intensity and duration (*75*). Pain prevalence, number of pain problems, and pain interference were reported.

***The International Spinal Cord Injury Quality of Life Basic Data Set (QoL-BDS) version 2.0.*** The QoL-BDS version 2.0 consists of four items for individuals to rate their satisfaction with life as a whole, physical health, psychological health, and social life (*76, 77*).

***Spinal Cord Independence Measure-Self Care (SCIM-SC).*** The SCIM III is the comprehensive ability rating scale for individuals with SCI that consists of three principal areas of daily living function: self-care, respiration and sphincter management, and mobility (*78, 79*). This study specifically focused on the self-care subscore, which includes feeding, bathing, dressing, and grooming.

***Modified Ashworth Scale (MAS).*** The MAS was used to evaluate muscle spasticity by rating muscle resistance on a 0–4 ordinal scale (*80*). Assessments were performed before each BCI-ESCS session on the right upper limb, targeting the shoulder flexor muscles and both extensor and flexor muscle groups of the elbow, wrist, and fingers. These repeated evaluations enabled close monitoring of potential spasticity changes related to the longitudinal intervention, with spasticity remaining stable and no evidence of increased muscle tone (Fig. S7).

***Nine-Hole Peg Test (9HPT).*** The 9HPT was used to assess finger dexterity following neurological injury (*81, 82*). The participant was instructed to pick up the pegs one at a time and place them in the holes as quickly and accurately as possible. Since the participant could not complete the test, the number of pegs successfully placed within 50 seconds was counted.

***Box and Block Test (BBT).*** The BBT measures manual dexterity by counting the number of wooden blocks (2.5 cm each) that a participant can transport from one side of a box to the other within 60 seconds, typically in individuals with central paresis (*83, 84*). Participants are instructed to place their hand on the table beside the box. Upon the examiner's cue to begin, they reach into the box and move one block at a time across a central partition to the opposite side, repeating the action as many times as possible within the allotted time. Only blocks that fully cross the divider are counted toward the final score, and only one block may be moved at a time.

**Maximum grip assessment**

Grip force using a hand dynamometer (Model: 12-0021, Baseline) and volitional EMG activity were assessed during a 5-second maximum voluntary contraction (MVC) task, in which the participant was instructed to grip as strongly as possible, with two trials per condition. EMG signals were recorded from the FCR and ECR muscles, and root-mean-square (RMS) EMG amplitude was calculated after applying a notch filter at the stimulation frequency to attenuate stimulation-related artifacts (*13*). Peak grip force and RMS EMG amplitude were compared to assess the immediate assistive effects across no stimulation, Tonic-ESCS, and BCI-ESCS conditions.

**3D reaching assessment**

***Reaching task***. The participant was asked to reach forward to four targets (center, left, right, and upward targets) with varying heights and lateral positions. The participant was seated in his wheelchair with the seat angle at 0° and seat-to-backrest at 90°. The targets were positioned on an adjustable-height table 20 cm above the participant's knees. The center target was positioned 40 cm horizontally from the body, aligned with the right shoulder. Left and right targets were horizontally placed 20 cm to the left and right of the center, and the upward target was placed vertically 20 cm above the center. A single reaching cycle was defined as the hand moving from the initial position to the target (reaching phase) and returning to the initial position (pull phase). The reaching task was repeated eight times per target before the next target was assigned in randomized order. The participant was instructed to reach as accurately as possible at a self-selected pace without time constraints. These reaching movements differed from those used during the training sessions, minimizing the likelihood of task learning effects. We compared reaching kinematics and muscle synergies before and after four weeks of BCI-ESCS use (longitudinal effects), and during the use of BCI-ESCS and Tonic-ESCS (assistive effects).

***Kinematics.*** Marker trajectory data of right-hand movement were acquired and sampled at 100 Hz using a twelve-camera motion capture system (Vicon Vero, Oxford, UK). One reflective marker was placed on each of the four targets, and another marker was placed at the ulnar styloid to track the wrist trajectory. The kinematic analysis focused on reaching trajectories, velocity peaks, target error, and initial direction angle for each target. All kinematic computations were performed in 3D using the full marker trajectories. For visualization, reaching trajectories were plotted in 2D planes according to target direction: the horizontal plane for the center, left, and right targets, and the vertical plane for the upward target (Figs. 2C and 4D). For the kinematic analysis, data from all four target directions were pooled to compare the overall effects of stimulation condition on reaching performance (Figs. 2C and 4D; Tables S1 and S7).

Velocity peaks were analyzed as a measure of movement smoothness, which is derived directly from the velocity profile and minimizes sensitivity to high-frequency measurement noise (*13*). This metric quantified the total number of local maxima and minima in the velocity profile during both the reaching and pulling phases. Movements with a higher number of velocity peaks indicated less smooth movement, characterized by frequent accelerations and decelerations (*13*). To identify these peaks, we applied the *findpeaks* function in MATLAB with a prominence threshold of 10% of the velocity profile's range. This threshold filtered out minor noise and ensured that only significant variations in velocity were captured. Target error was calculated as the 3D Euclidean distance between the wrist marker position and the corresponding target position at reaching termination. The initial direction angle was calculated as the angular deviation between the vector from the onset position to the target and the vector from the onset position to the hand position at the end of the initial movement phase, defined as the point of maximum velocity (*85, 86*). The calculation was performed in MATLAB using the *acos* function to determine the angle in radians, which was then converted to degrees, ensuring an objective assessment of the initial movement direction relative to the intended trajectory.

***Muscle synergy.*** We quantified muscle synergies to characterize the coordination-level organization during reaching (Figs. S4 and S6), providing a descriptive summary of how patterns of muscle co-activation and their temporal recruitment relate to reaching control. Coordinated movements, such as upper limb reaching, rely on the precise and timely coactivation of multiple muscles to ensure accurate and controlled limb motion. Muscle synergies were extracted using nonnegative matrix factorization (NNMF), a dimensionality reduction technique to investigate the muscle coordination patterns underlying these movements (*13*). Electromyography (EMG) data were collected during the reaching task, capturing muscle activity from multiple upper-limb muscles.

EMG activity was recorded using a wireless EMG system (Trigno Wireless System; Delsys, Boston, MA). The EMG signals were band-pass filtered (20–450 Hz) and sampled at 2,148 Hz. Sensors were attached to the following muscles on the right side of the body: infraspinatus (IF), supraspinatus (SSP), lower trapezius (LT), middle trapezius (MT), upper trapezius (UT), deltoid anterior (DA), deltoid middle (DM), deltoid posterior (DP), serratus anterior (SA), pectoralis major clavicular head (PC), biceps brachii long head (BCL), triceps brachii long head (TLO), brachioradialis (BR), pronator teres (PT), flexor carpi ulnaris (FCU), and extensor carpi radialis (ECR).

In post-processing, EMG signals were high-pass filtered at 40 Hz, rectified, low-pass filtered at 10 Hz, and normalized to the average of the maximum EMG values across all trials for each muscle (*87*). Trials were concatenated sequentially to form a $m \times t$ data matrix, where $m$ represents the number of muscles (16 muscle groups), and $t$ represents the total number of time points, calculated as the product of trials and time bins. Each trial included 100-time bins for the reaching phase and 100-time bins for the pulling phase, totaling 200-time bins. Each of the four targets was repeated for eight trials, resulting in a total of 32 trials. Thus, the complete dataset consisted of $t = 6{,}400$ time bins (32 trials × 200 bins per trial).

Muscle synergies were identified from the EMG matrices ($M$) using NNMF, which decomposes the EMG signals into two components: a synergy vector, representing the contribution of each muscle to the synergy, and a temporal activation pattern, describing the time course of each synergy during the movement, as shown in the following equation:

$$M = W \cdot C + e$$

where $M$ ($m \times t$ matrix) is a linear combination of a synergy vector $W$ ($m \times n$ matrix, where $n$ is the number of muscle synergies) and $C$ ($n \times t$ matrix, representing temporal activation patterns), and $e$ is the residual error matrix. To determine the number of muscle synergies, NNMF was applied to extract each possible synergy from 1 to 16 for each dataset, and the variance accounted for (VAF) by the reconstructed EMG was computed for each number of synergies tested (*88*). Each synergy extraction was repeated 50 times with randomized initialization to avoid local minima, and the iteration with the highest VAF was selected (*33*, *87*, *89–91*).

The contributions of individual muscles to each synergy were quantified using the synergy vector. Temporal activation patterns of the synergies were analyzed to examine the timing and amplitude of muscle coordination during the task. All repetitions of the task were included in the analysis to ensure robust and representative results.

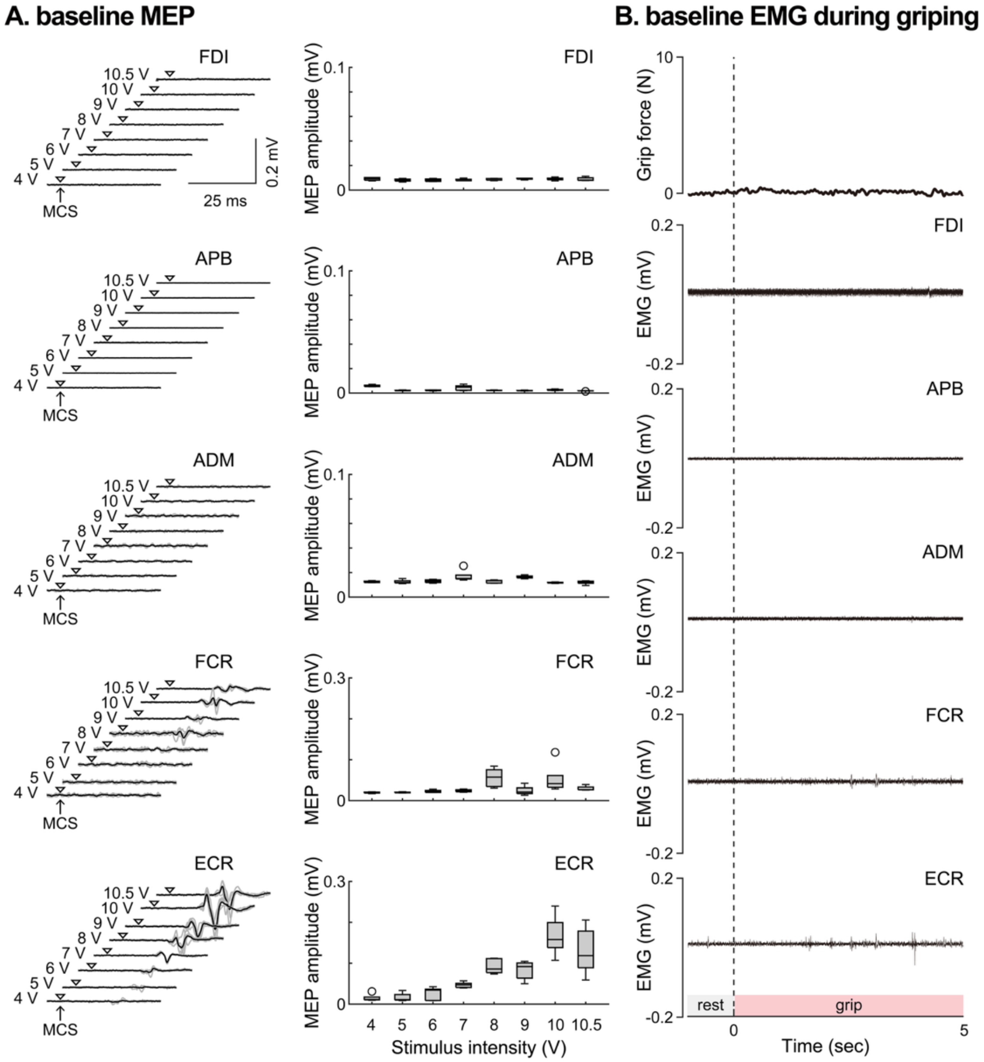


**Fig. S1. Baseline neurophysiological evidence supporting motor-complete classification.**
**(A)** Baseline motor-evoked potentials (MEPs) (5 stimuli per intensity). Representative MEP waveforms and boxplots of peak-to-peak MEP amplitudes evoked by single-pulse motor cortex stimulation (MCS) across stimulation intensities (4–10.5 V) in intrinsic hand muscles (first dorsal interosseous, FDI; abductor pollicis brevis, APB; abductor digiti minimi, ADM) and forearm muscles (flexor carpi radialis, FCR; extensor carpi radialis, ECR). At baseline, no measurable MEPs were observed in the intrinsic hand muscles across intensities, whereas small responses were detectable in forearm muscles. **(B)** Baseline surface EMG during grip attempt. Representative EMG traces recorded during a grip attempt at baseline, showing no observable voluntary EMG activities in intrinsic hand muscles (FDI, APB, ADM). Forearm recordings (FCR, ECR) are shown for comparison. The shaded bar indicates the grip epoch. The presence of responses in forearm muscles, alongside the absence of baseline MEPs and volitional EMG in intrinsic hand muscles (innervated below C5), supports the classification of the participant as having motor-complete SCI at the C4–C5 level.

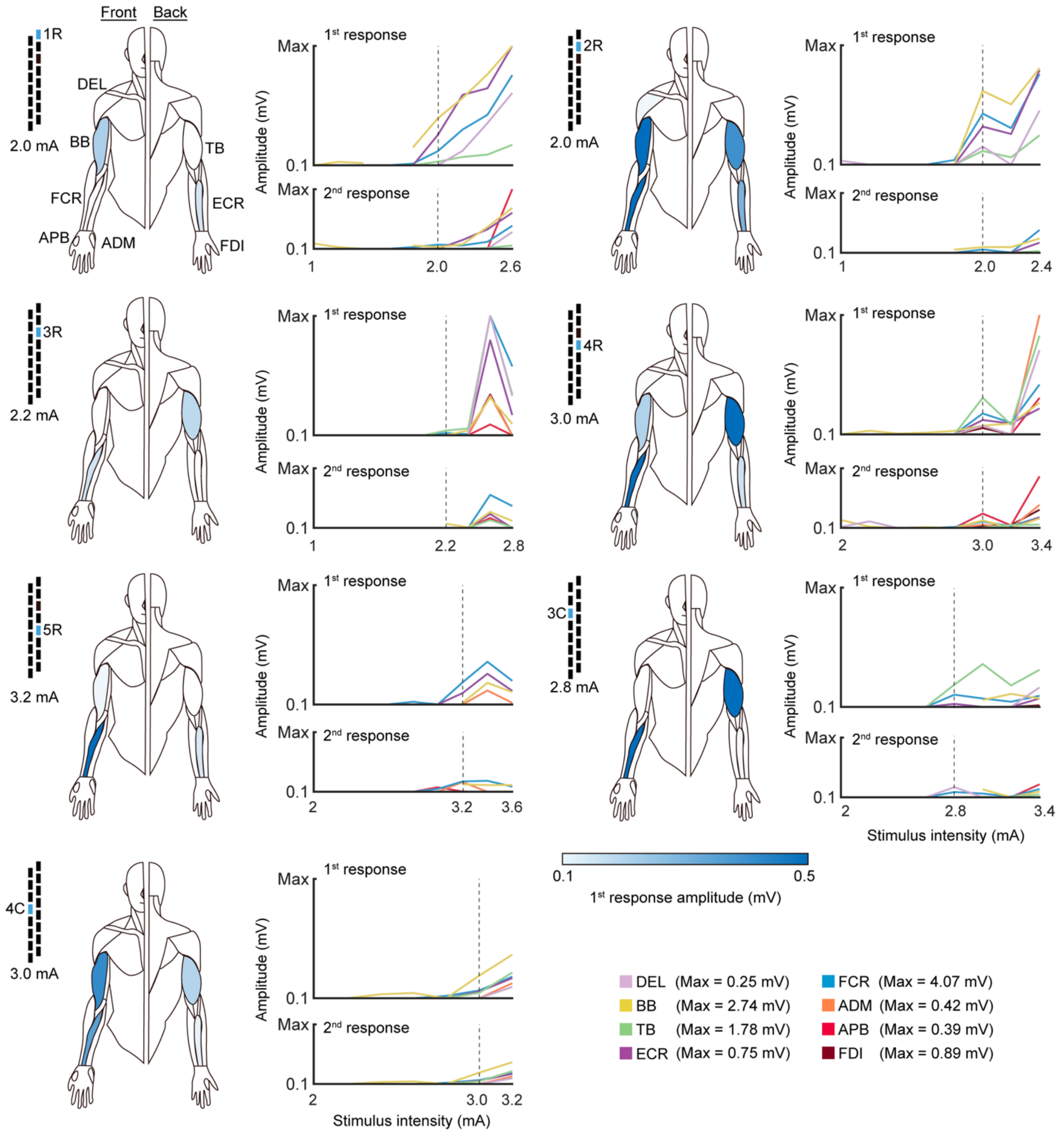


**Fig. S2. Spinal mapping for optimizing ESCS contacts.**
Spinal mapping results for optimizing epidural spinal cord stimulation (ESCS) contacts. Stimulation was applied through different contacts along the epidural electrode array, and muscle responses were recorded from multiple upper-limb muscles, including the deltoid (DEL), biceps brachii (BB), triceps brachii (TB), extensor carpi radialis (ECR), flexor carpi radialis (FCR), abductor digiti minimi (ADM), abductor pollicis brevis (APB), and first dorsal interosseous (FDI). Paired-pulse ESCS with incremental stimulation intensities (3 pulses per intensity) was used to assess the contact-specific recruitment curves and to confirm that stimulation predominantly engaged the dorsal roots rather than directly activating motor axons, as indicated by suppression of the second response relative to the first. The human figurines show predominant muscle responses, based on first-response amplitude, at stimulation intensities selected from the ascending portion of recruitment curves to illustrate contact-specific recruitment patterns. The recruitment curves show peak-to-peak amplitudes of the first and second responses across increasing stimulation intensities. Y-axes are scaled to each muscle's maximum response across all contacts and intensities. Vertical dotted lines indicate the stimulation intensities corresponding to the human figurine representations. Truncated portions of the recruitment curves indicate missing data points caused by spastic responses, particularly in the BB muscle. These spinal mapping results were used to determine the optimal ESCS contacts for neuromodulation and motor engagement during BCI-ESCS therapy.

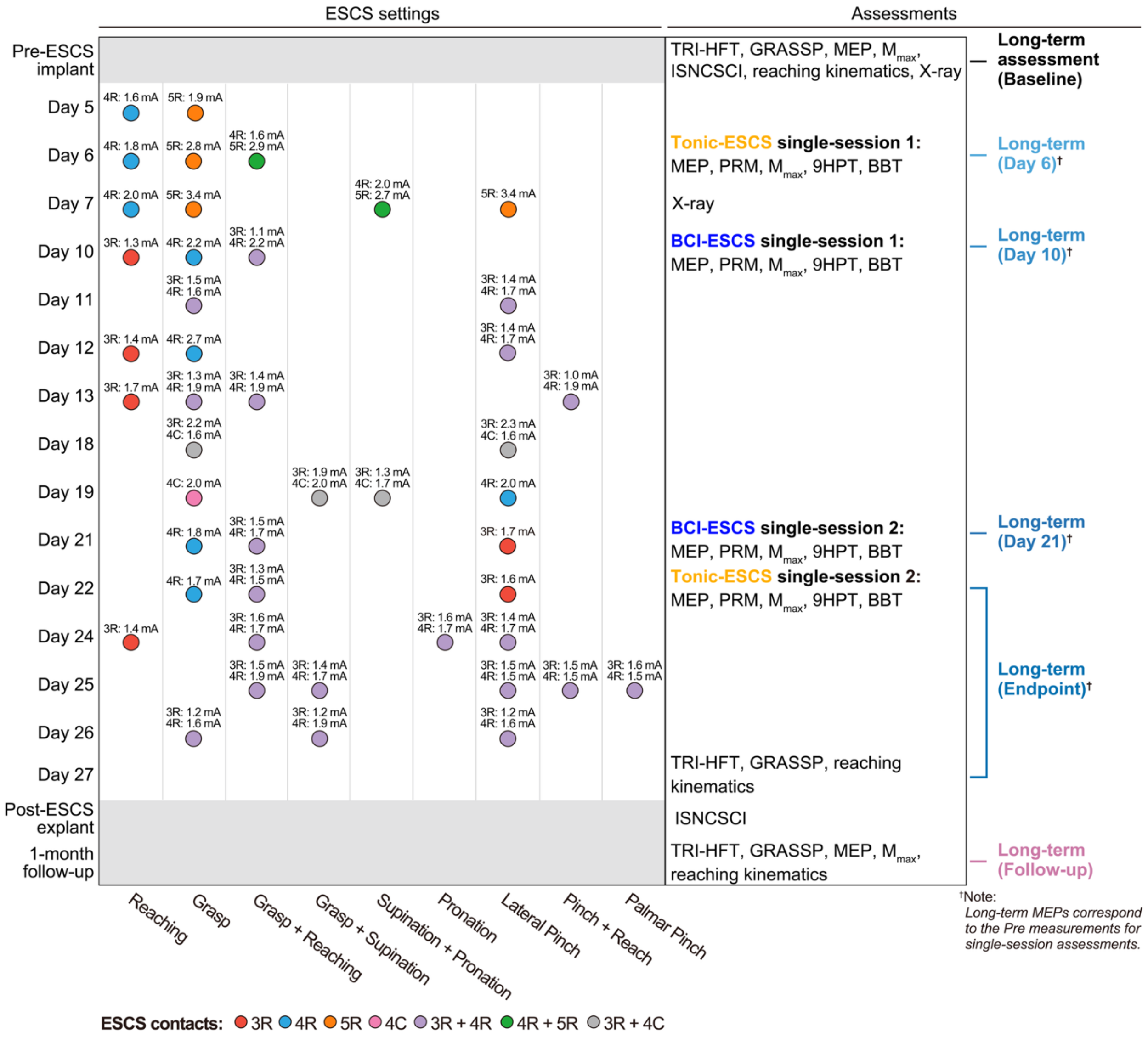


**Fig. S3. Timeline of stimulation parameters and assessments throughout BCI-ESCS therapy.**

Color-coded markers indicate the stimulation contacts and intensities applied for each motor task across sessions, with all stimulation delivered at 80 Hz. At the beginning of each therapy session, stimulation contacts and intensities were adjusted to optimize neuromodulation for each movement task. X-ray imaging was performed at Baseline and Day 7 to confirm lead placement and re-optimize stimulation parameters. Assessments conducted throughout the intervention are listed on the right. Single-session assessments of BCI-ESCS and Tonic-ESCS included neurophysiological (motor evoked potentials (MEP), posterior root muscle (PRM) reflex, and maximum M-wave ($M_{max}$)) and functional (Nine-Hole Peg Test (9HPT) and Box and Block Test (BBT)) assessments. Longitudinal assessments included Toronto Rehabilitation Institute-Hand Function Test (TRI-HFT), Graded Redefined Assessment of Strength, Sensibility, and Prehension (GRASSP), and reaching kinematics at Baseline, Endpoint, and Follow-up; International Standards for Neurological Classification of Spinal Cord Injury (ISNCSCI) examinations at Baseline and Endpoint; and MEP assessments at Baseline, Day 6, Day 10, Day 21, Endpoint (Day 22), and Follow-up. The MEP recordings obtained at Day 6, Day 10, Day 21, and Day 22 also corresponded to the Pre measurements for the respective single-session BCI-ESCS or Tonic-ESCS assessment.

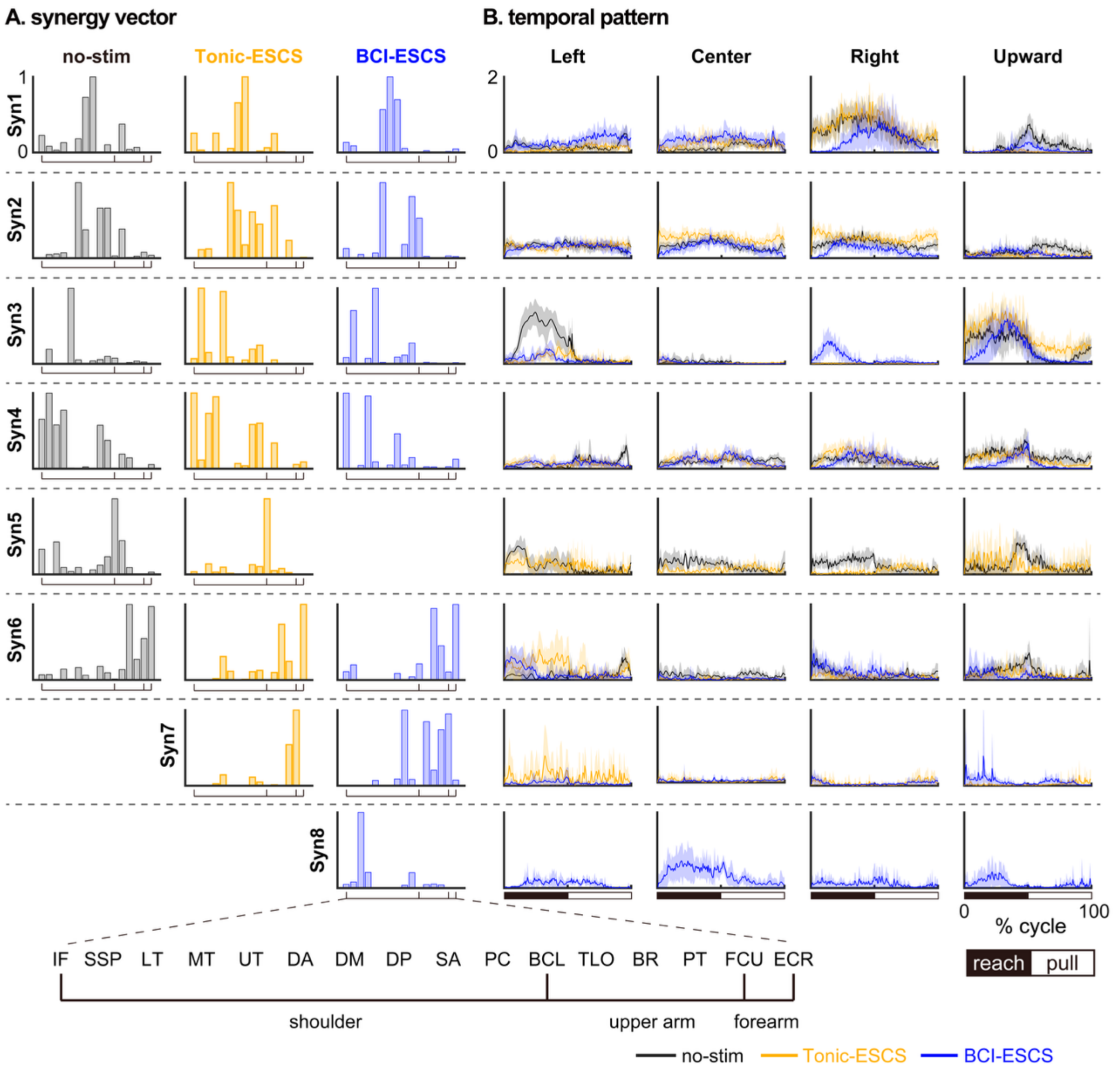


**Fig. S4. Assistive effects of BCI-ESCS and Tonic-ESCS on muscle synergies.**
Muscle synergies were extracted under three conditions: no stimulation, Tonic-ESCS, and BCI-ESCS. **(A)** Synergy vectors showing muscle contributions. Both stimulation modes increased the number of synergies to seven compared to six in no stimulation, suggesting a shift from co-activation toward more differentiated coordination patterns. Shoulder-related synergies reorganized from broad patterns (Syn3: UT; Syn4: IF, SSP, LT, MT) into more joint-function-related synergies under stimulation (Syn3: SSP, UT for elevation; Syn4: IF, MT for external rotation). Similarly, wrist-related synergies differentiated from a single co-activated pattern (Syn6: BR, PT, FCU, ECR) into two distinct synergies with stimulation (Syn6: BR, ECR for extension; Syn7: FCU, PT for flexion/pronation). BCI-ESCS particularly exhibited an LT-dominant synergy (Syn8), whereas Tonic-ESCS retained the BCL-dominant synergy (Syn5) similar to the no-stimulation condition, suggesting that BCI-triggered stimulation may facilitate scapular stabilizer activation and reduced reliance on the elbow flexor-dominant coordination. **(B)** Temporal activation patterns during reaching showed target-dependent features. Under both stimulation modes, direction-specific burst patterns, such as the UT-biased burst in Syn3 toward the Left target, were attenuated, with shoulder activity redistributed across multiple synergies. This suggests that stimulation altered temporal coordination patterns rather than relying on a single dominant burst.
**Muscle abbreviations**: BCL: biceps long head, BR: brachioradialis, DA: anterior deltoid, DM: middle deltoid, DP: posterior deltoid, ECR: extensor carpi radialis longus, FCU: flexor carpi ulnaris, IF: infraspinatus, LT: lower trapezius, MT: middle trapezius, UT: upper trapezius, PC: pectoralis, PT: pronator teres, SA: serratus anterior, SSP: supraspinatus, TLO: triceps long head.

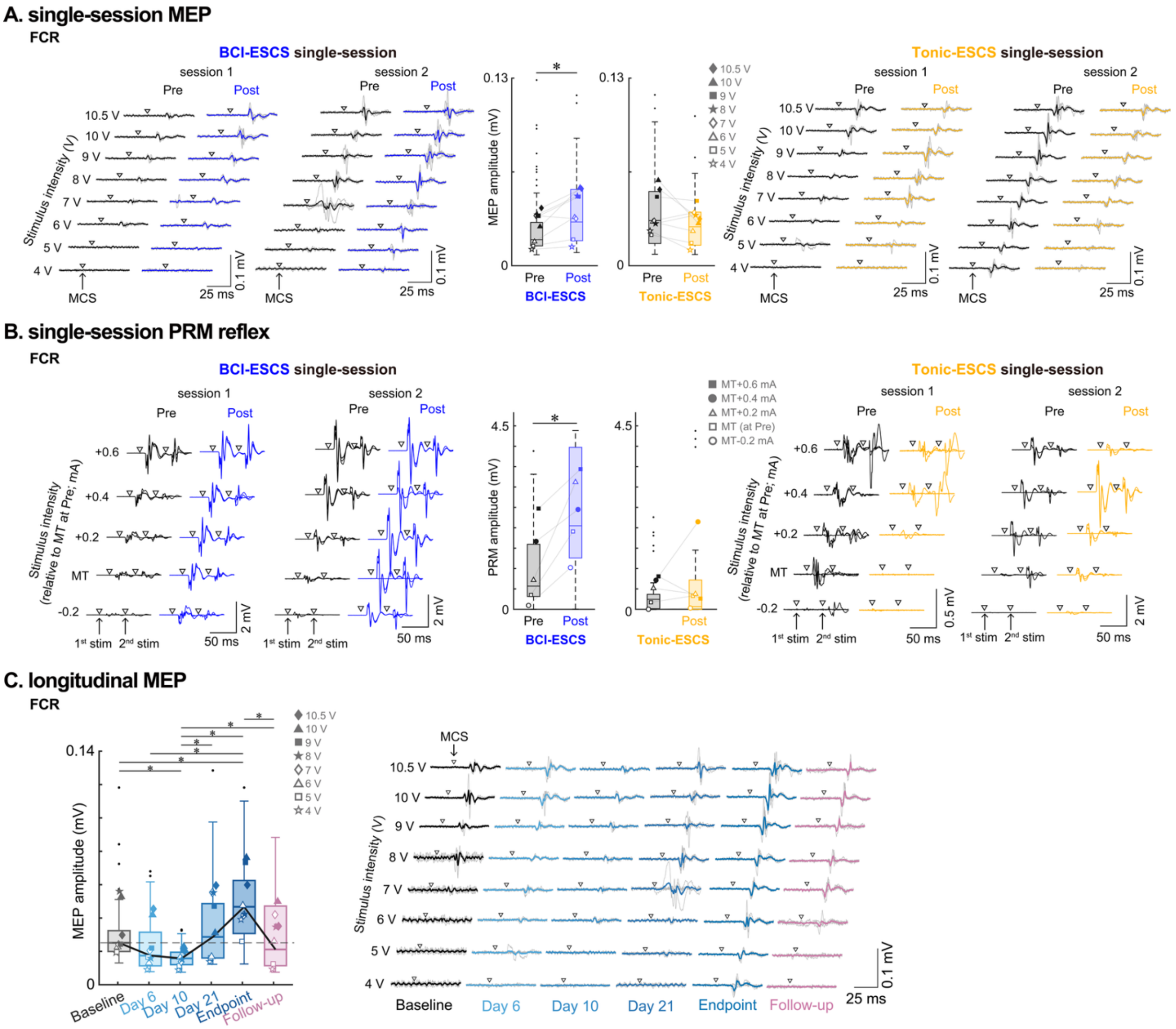


**Fig. S5. Neurophysiological responses in the FCR muscle.**

**(A)** Single-session motor evoked potential (MEP) responses following BCI-ESCS and Tonic-ESCS use. MEPs were recorded across 4–10.5 V stimulation intensities (8 intensities × 5 trials per session), with waveforms from both sessions shown for each condition. Boxplots show peak-to-peak MEP amplitudes pooled across stimulation intensities and sessions, with individual symbols representing the mean response at each intensity. **(B)** Single-session posterior root muscle (PRM) reflex responses following BCI-ESCS and Tonic-ESCS use. PRM reflexes were elicited across stimulation intensities relative to Pre-measurement motor threshold (MT), from MT−0.2 to MT+0.6 mA (5 intensities × 3 trials per session), with waveforms from both sessions shown for each condition. Boxplots show peak-to-peak PRM reflex amplitudes pooled across stimulation intensities and sessions. **(C)** Longitudinal MEP responses across six timepoints. Boxplots show peak-to-peak MEP amplitudes pooled across stimulation intensities at each assessment timepoint (8 intensities × 5 trials per assessment timepoint), with waveforms shown across all six assessment timepoints at each tested stimulus intensity.

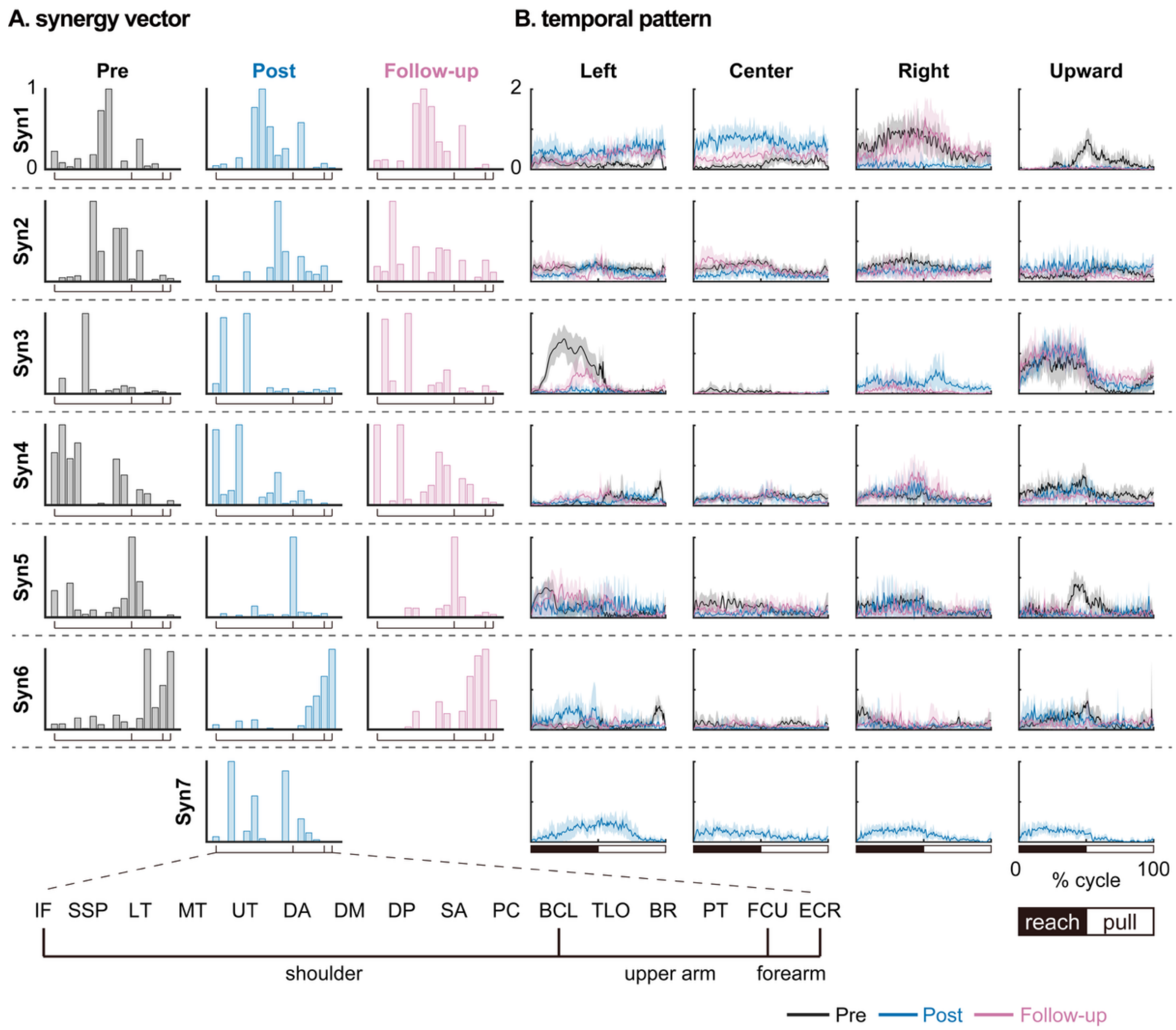


**Fig. S6. Longitudinal effects of BCI-ESCS use on muscle synergies.**
Muscle synergies were extracted before therapy (Baseline), after four weeks of BCI-ESCS therapy (Endpoint), and one month after lead explantation (Follow-up). **(A)** Synergy vectors showing muscle contributions. BCI-ESCS therapy increased the number of muscle synergies after four weeks of BCI-ESCS use in the absence of stimulation. Shoulder-related synergies at Baseline (Syn3, Syn4) showed broad co-activation patterns, whereas at Endpoint they became functionally organized, with Syn3 engaging SSP and UT for shoulder elevation, and Syn4 engaging IF and MT for external rotation. Additionally, an LT-dominant synergy (Syn7) emerged at Endpoint across all reaching directions, consistent with the improved scapular coordination. **(B)** Temporal activation patterns during reaching. At Endpoint, shoulder activity became more evenly distributed across synergies. During the center-target task, Syn1 engagement increased, reflecting greater recruitment of DA, DM, and DP. During the upward-target task, Syn1 contribution decreased, and a broader repertoire of shoulder synergies emerged, including Syn7, resulting in more balanced activation timing across modules. At Follow-up, Syn7 was no longer present, potentially explaining the reversion of certain kinematic outcomes toward baseline. **Muscle abbreviations:** BCL: biceps long head, BR: brachioradialis, DA: anterior deltoid, DM: middle deltoid, DP: posterior deltoid, ECR: extensor carpi radialis longus, FCU: flexor carpi ulnaris, IF: infraspinatus, LT: lower trapezius, MT: middle trapezius, UT: upper trapezius, PC: pectoralis, PT: pronator teres, SA: serratus anterior, SSP: supraspinatus, TLO: triceps long head.

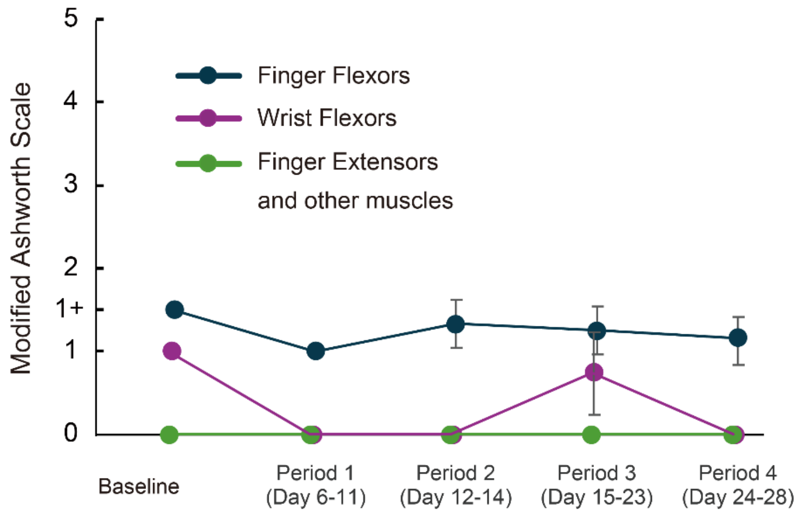


**Fig. S7. Longitudinal assessment of upper limb spasticity using the Modified Ashworth Scale (MAS).**
Spasticity was evaluated using the Modified Ashworth Scale (MAS), which rates muscle resistance to passive movement on a 0–4 ordinal scale. Assessments were conducted before each epidural spinal cord stimulation (ESCS) therapy session on the right upper limb, targeting the shoulder flexor muscles and the flexor and extensor muscles of the elbow, wrist, and fingers. For visualization, MAS scores were averaged across sessions within five time periods: Baseline, Period 1 (Days 6–11), Period 2 (Days 12–14), Period 3 (Days 15–23), and Period 4 (Days 24–28). Spasticity remained stable throughout the intervention, with no evidence of increased muscle tone. Data are shown as mean ± standard deviation across evaluations for each muscle group.

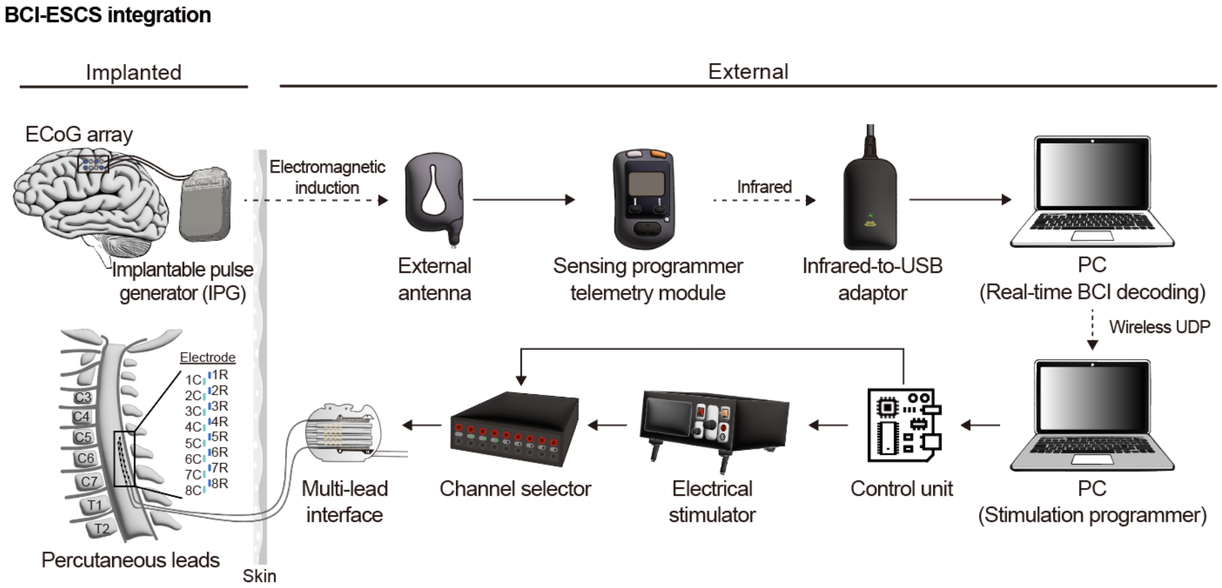


**Fig. S8. BCI-ESCS system integration.**
Schematic representation of the brain-computer interface (BCI)-controlled epidural spinal cord stimulation (ESCS) system, illustrating the interaction between implanted and external components. The implanted system consists of an electrocorticography (ECoG) array connected to an implantable pulse generator (IPG), which was used for ECoG signal acquisition. ESCS was delivered via an external constant-current stimulator. Separately, percutaneous epidural leads are implanted in the cervical spinal cord to deliver ESCS. Neural signals from the ECoG array are transmitted wirelessly via an external antenna through electromagnetic induction to a sensing programmer telemetry module, which relays the data to a PC for real-time BCI decoding. Upon detection of motor intention, a control unit processes the decoded signals and triggers the electrical stimulator via a channel selector, directing current through the percutaneous leads to selectively activate optimized channels. Stimulation parameters are managed through a separate PC interface, allowing precise control of pulse timing and amplitude. This implantable BCI-ESCS system enables real-time, intention-driven spinal modulation to restore volitional upper-limb movement in individuals with chronic spinal cord injury (SCI).

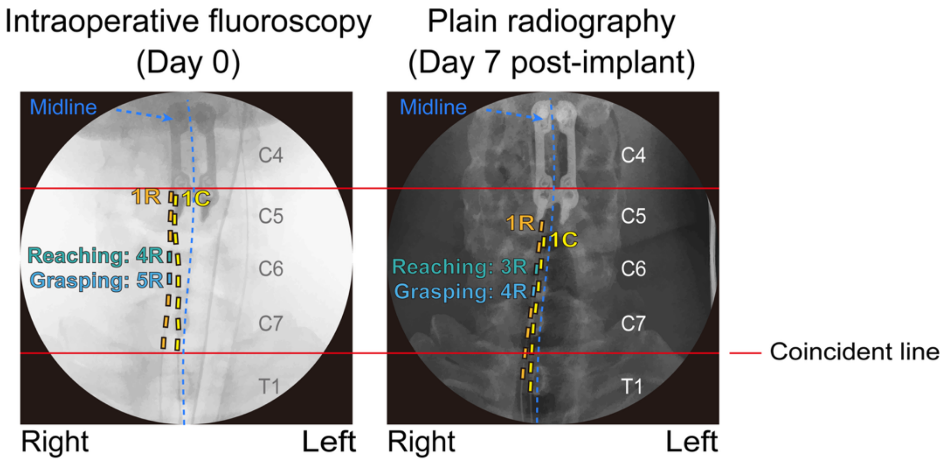


**Fig. S9. Epidural spinal cord stimulation (ESCS) lead position during early implantation period**.
X-ray images on day 0 (intraoperative fluoroscopy, originally anteroposterior, mirrored image) and day 7 (plain radiography, posteroanterior). Red lines mark consistent anatomical reference points across both X-ray images to assist interpretation. Minor displacement following initial implantation resulted in shifted optimal contacts for therapy (reaching: from 4R to 3R; grasping: from 5R to 4R). As subsequent spinal mapping after day 7 confirmed stable recruitment profiles throughout Weeks 2–4, no additional radiography was acquired during this period.

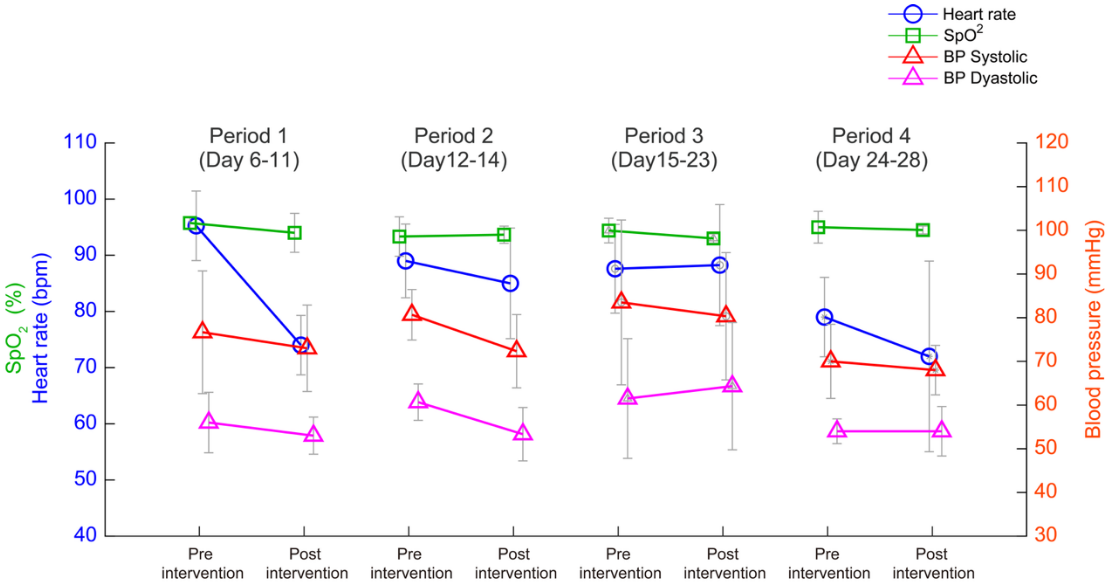


**Fig. S10. Stability of vital signs during the study period.**
Vital signs were measured before and after each BCI-ESCS therapy session and averaged across four time windows: Period 1 (Day 6–11), Period 2 (Day 12–14), Period 3 (Day 15–23), and Period 4 (Day 24–28). Shown are heart rate (blue), oxygen saturation ($SpO_2$; green), systolic blood pressure (BP; orange), and diastolic BP (magenta). Across all time points, vital signs remained stable. Data are shown as mean ± standard deviation.

**Table S1. Statistical results of assistive effects.** Comparisons of maximum voluntary contraction (MVC) and electromyography (EMG) of the flexor carpi radialis (FCR) and extensor carpi radialis (ECR) muscles during maximum grip (Fig. 2A) and 3D reaching kinematics (Fig. 2C) were performed between conditions (no-stim, Tonic-ESCS, and BCI-ESCS). For the MVC (2 attempts per condition), effect sizes (Cohen's d) are reported as descriptive indicators given the limited sample size. For the 3D reaching kinematics, comparisons of velocity peaks, target error, and initial direction angle were performed using a bootstrap method on data pooled across four reaching targets (left, right, upward, and center) (8 attempts × 4 targets per condition), with results reported as two-sided p-values with Bonferroni correction for three comparisons and effect sizes (Cohen's d). Asterisks indicate statistically significant differences.

| Assistive effects on grip maximum voluntary contraction (MVC) | | | | | | |
|---|---|---|---|---|---|---|
| Comparison | Grip Force | | FCR EMG | | ECR EMG | |
| | d | | d | | d | |
| no-stim − Tonic-ESCS | -13.030 | | -61.596 | | -6.009 | |
| no-stim − BCI-ESCS | -2.803 | | -22.149 | | -5.877 | |
| Tonic-ESCS − BCI-ESCS | -1.464 | | -9.404 | | -4.009 | |
| **Assistive effects on 3D reaching kinematics** | | | | | | |
| Comparison | Velocity peaks | | Target error | | Initial direction angle | |
| | p | d | p | d | p | d |
| no-stim − Tonic-ESCS | *<0.001 | 1.733 | *<0.001 | 0. 722 | 0.184 | -0.376 |
| no-stim − BCI-ESCS | *<0.001 | 1.225 | *<0.001 | 1.101 | *<0.001 | -1.654 |
| Tonic-ESCS − BCI-ESCS | *0.002 | -0.870 | *<0.001 | 0.964 | *<0.001 | -1.574 |

**Table S2. Statistical results of single-session effects on corticospinal and spinal excitability.** Motor-evoked potential (MEP) responses (8 intensities × 5 trials per session) and posterior root muscle (PRM) reflexes (5 intensities × 3 trials per session) in the extensor carpi radialis (ECR) and flexor carpi radialis (FCR) muscles (Figs. 3B-C and S5A-B) were compared between Pre and Post assessments for each condition using a bootstrap method. It should be noted that ECR MEP and PRM reflex amplitudes were normalized to the corresponding maximum M-wave ($M_{max}$) values. Data were pooled across all stimulation intensities and across two sessions within each condition. The table reports all pairwise comparisons among four groups (BCI-ESCS Pre, BCI-ESCS Post, Tonic-ESCS Pre, Tonic-ESCS Post). Results are reported as two-sided p-values with Bonferroni correction for six comparisons and effect sizes (Cohen's d). Asterisks indicate statistically significant differences.

| Single-session effects on motor-evoked potentials (MEP) | | | | |
|---|---|---|---|---|
| Pairwise comparison | ECR muscle (normalized to $M_{max}$) | | FCR muscle | |
| | p | d | p | d |
| BCI-ESCS pre − BCI-ESCS post | *0.005 | -0.483 | *0.023 | -0.465 |
| Tonic-ESCS pre − Tonic-ESCS post | >0.999 | 0.083 | 0.074 | 0.386 |
| BCI-ESCS pre − Tonic-ESCS pre | 0.590 | -0.254 | 0.056 | -0.433 |
| BCI-ESCS post − Tonic-ESCS post | 0.419 | 0.287 | *0.040 | 0.421 |
| BCI-ESCS pre − Tonic-ESCS post | >0.999 | -0.159 | >0.999 | -0.101 |
| BCI-ESCS post − Tonic-ESCS pre | >0.999 | 0.206 | >0.999 | 0.026 |
| **Single-session effects on posterior root muscle (PRM) reflexes** | | | | |
| Pairwise comparison | ECR muscle (normalized to $M_{max}$) | | FCR muscle | |
| | p | d | p | d |
| BCI-ESCS pre − BCI-ESCS post | *0.012 | -0.732 | *<0.001 | -1.078 |
| Tonic-ESCS pre − Tonic-ESCS post | >0.999 | -0.015 | >0.999 | -0.188 |
| BCI-ESCS pre − Tonic-ESCS pre | *<0.001 | 1.347 | *0.025 | 0.696 |
| BCI-ESCS post − Tonic-ESCS post | *<0.001 | 2.173 | *<0.001 | 1.339 |
| BCI-ESCS pre − Tonic-ESCS post | *<0.001 | 1.257 | 0.913 | 0.368 |
| BCI-ESCS post − Tonic-ESCS pre | *<0.001 | 2.341 | *<0.001 | 1.819 |

**Table S3. Longitudinal effects of BCI-ESCS therapy on the Toronto Rehabilitation Institute–Hand Function Test (TRI-HFT) 10-object manipulation and rectangular block scores**. The assessments were conducted at Baseline, Endpoint, and Follow-up.

| Object manipulation | | Baseline | | Endpoint | | Follow-up | |
|---|---|---|---|---|---|---|---|
| | | Right | Left | Right | Left | Right | Left |
| Ten objects | | | | | | | |
| 1. Mug | | 4 | 4 | 4 | 4 | 4 | 4 |
| 2. Paper | | 1 | 1 | 2 | 1 | 4 | 1 |
| 3. Book | | 1 | 1 | 1 | 1 | 1 | 1 |
| 4. Ziplock bag | | 2 | 2 | 2 | 2 | 2 | 2 |
| 5. Pop can | | 1 | 2 | 2 | 2 | 2 | 2 |
| 6. Dice | | 4 | 2 | 4 | 2 | 4 | 2 |
| 7. Sponge | | 1 | 1 | 1 | 1 | 1 | 1 |
| 8. Credit card | | 1 | 1 | 4 | 1 | 4 | 4 |
| 9. Mobile phone | | 2 | 2 | 6 | 6 | 6 | 4 |
| 10. Pencil | | 2 | 2 | 2 | 2 | 2 | 2 |
| Subtotal (70) | | 19 | 18 | 28 | 22 | 30 | 23 |
| Subtotal Right+Left (140) | | 37 | | 50 | | 53 | |
| Rectangular blocks | | | | | | | |
| Weight | Surface friction level | | | | | | |
| 100 g | high | 4 | 4 | 4 | 4 | 4 | 4 |
| | medium | 1 | 4 | 4 | 4 | 4 | 4 |
| | low | 2 | 4 | 4 | 4 | 4 | 4 |
| 200 g | high | 2 | 4 | 4 | 4 | 4 | 4 |
| | medium | 2 | 4 | 4 | 4 | 4 | 4 |
| | low | 2 | 2 | 4 | 4 | 4 | 4 |
| 300 g | high | 2 | 2 | 4 | 4 | 4 | 4 |
| | medium | 2 | 2 | 4 | 4 | 4 | 4 |
| | low | 2 | 2 | 4 | 2 | 4 | 2 |
| Subtotal (63) | | 19 | 28 | 36 | 34 | 36 | 34 |
| Subtotal Right+Left (126) | | 47 | | 70 | | 70 | |

**Table S4. Statistical results of longitudinal effects on the Toronto Rehabilitation Institute–Hand Function Test (TRI-HFT) 10-object manipulation and rectangular block scores.** Comparison of TRI-HFT scores in the 10-object manipulation and rectangular block tasks (Fig. 4B) between the study participant (n=1) and a control cohort (n=6) who underwent conventional therapy only. Results are reported as two-sided p-values with Bonferroni correction for multiple comparisons, and effect sizes (Cohen's d). Asterisks indicate significant differences.

| Comparison between the study participant and a control cohort on TRI-HFT | | |
|---|---|---|
| TRI-HFT task | p | d |
| 10-object manipulation | *<0.001 | 1.160 |
| Rectangular blocks | *0.001 | 1.187 |

**Table S5. Longitudinal effects of BCI-ESCS therapy on the Graded Redefined Assessment of Strength, Sensibility and Prehension (GRASSP) scores**. The assessments were conducted at Baseline, Endpoint, and Follow-up.

| | Baseline | | Endpoint | | Follow-up | |
|---|---|---|---|---|---|---|
| | Right | Left | Right | Left | Right | Left |
| Strength | 7 | 10 | 8 | 9 | 8 | 9 |
| Sensory-Dorsal | 0 | 0 | 0 | 3 | 0 | 1 |
| Sensory-Palmar | 1 | 4 | 2 | 0 | 0 | 0 |
| Prehension-Qualitative | 2 | 2 | 2 | 1 | 2 | 1 |
| Prehension-Quantitative | 2 | 6 | 6 | 5 | 5 | 5 |
| Total | 12 | 22 | 18 | 18 | 15 | 16 |

**Table S6. Longitudinal effects of BCI-ESCS therapy on the International Standards for Neurological Classification of Spinal Cord Injury (ISNCSCI) upper extremity motor scores (UEMS) and zones of partial preservation (ZPP) assessments.** The assessments were conducted at Baseline and Endpoint.

| | Baseline | | Endpoint | |
|---|---|---|---|---|
| | Right | Left | Right | Left |
| Motor | | | | |
| C5 Elbow flexors | 4 | 4 | 4 | 5 |
| C6 Wrist extensors | 0 | 0 | 0 | 1 |
| C7 Elbow extensors | 0 | 0 | 2 | 2 |
| C8 Finger flexors | 0 | 0 | 0 | 0 |
| T1 Finger abductors | 0 | 0 | 0 | 1 |
| Total | 4 | 4 | 6 | 9 |
| Zone of partial preservation | | | | |
| Sensory | T9 | T10 | T10 | T11 |
| Motor | C5 | C5 | C7 | T1 |

**Table S7. Statistical results of longitudinal effects on reaching kinematics and corticospinal excitability.** Velocity peaks, target error, and initial direction angle during the 3D reaching kinematics task (Fig. 4D) were compared across Baseline, Endpoint, and Follow-up using a bootstrap method on data pooled across four reaching directions (left, right, upward, and center) (8 attempts × 4 targets per assessment time point). Motor-evoked potential (MEP) responses in the flexor carpi radialis (FCR) and extensor carpi radialis (ECR) muscles (Fig. 4E) were compared across Baseline, Day 6, Day 10, Day 21, Endpoint, and Follow-up using a bootstrap method on data pooled across all tested stimulation intensities at each timepoint (8 intensities × 5 trials per assessment time point) (Fig. S3). The table reports all pairwise comparisons between timepoints. Results are reported as two-sided p-values with Bonferroni correction and effect sizes (Cohen's d). Asterisks indicate statistically significant differences.

| Longitudinal effects on 3D reaching kinematics | | | | | | |
|---|---|---|---|---|---|---|
| Comparison | Velocity peaks | | Target error | | Initial direction angle | |
| | p | d | p | d | p | d |
| Baseline – Endpoint | *<0.001 | 1.512 | *<0.001 | 1.097 | 0.196 | -0.384 |
| Baseline – Follow-up | 0.848 | 0.243 | *<0.001 | 0.970 | *<0.001 | -0.964 |
| Endpoint – Follow-up | *<0.001 | -1.378 | 0.434 | -0.360 | *0.005 | -0.631 |

| Longitudinal effects on motor-evoked potentials (MEP) | | | | |
|---|---|---|---|---|
| Pairwise comparison | ECR muscle (normalized to $M_{max}$) | | FCR muscle | |
| | p | d | p | d |
| Baseline – Day 6 | >0.999 | -0.176 | 0.468 | 0.444 |
| Baseline – Day 10 | *0.003 | -0.763 | *0.002 | 0.989 |
| Baseline – Day 21 | >0.999 | -0.355 | >0.999 | -0.242 |
| Baseline – Endpoint | *0.002 | -1.133 | *0.003 | -0.848 |
| Baseline – Follow-up | >0.999 | -0.184 | >0.999 | 0.025 |
| Day 6 – Day 10 | 0.093 | -0.597 | 0.093 | 0.570 |
| Day 6 – Day 21 | >0.999 | -0.158 | 0.066 | -0.624 |
| Day 6 – Endpoint | *0.002 | -1.009 | *0.002 | -1.298 |
| Day 6 – Follow-up | >0.999 | -0.009 | >0.999 | -0.399 |
| Day 10 – Day 21 | 0.387 | 0.495 | *0.002 | -1.045 |
| Day 10 – Endpoint | 0.156 | -0.536 | *0.002 | -1.835 |
| Day 10 – Follow-up | 0.099 | 0.584 | *0.002 | -0.900 |
| Day 21 – Endpoint | *0.002 | -0.942 | 0.237 | -0.523 |
| Day 21 – Follow-up | >0.999 | 0.146 | >0.999 | 0.257 |
| Endpoint – Follow-up | *0.002 | 0.999 | *0.006 | 0.848 |

**Table S8. Longitudinal effects of BCI-ESCS therapy on the Spinal Cord Independence Measure-Self Care (SCIM-SC) scores**. The assessments were conducted at Baseline, Endpoint, and Follow-up.

| | | Baseline | Endpoint | Follow-up |
|---|---|---|---|---|
| Feeding | | 2 | 2 | 2 |
| Bathing | Upper body | 1 | 1 | 1 |
| | Lower body | 0 | 0 | 0 |
| Dressing | Upper body | 0 | 0 | 0 |
| | Lower body | 0 | 0 | 0 |
| Grooming | | 1 | 1 | 1 |
| Total | | 4 | 4 | 4 |

**Table S9. Longitudinal effects of BCI-ESCS therapy, International Spinal Cord Injury Quality of Life Basic Data Set (QoL-BDS) scores.** The assessments were conducted at Baseline, Endpoint, and Follow-up.

| | Baseline | Endpoint | Follow-up |
|---|---|---|---|
| Life as a whole | 9 | 9 | 9 |
| Physical health | 8 | 8 | 9 |
| Psychological health | 9 | 10 | 10 |
| Social life | 9 | 10 | 10 |

**Table S10. Longitudinal effects of BCI-ESCS therapy, International Spinal Cord Injury Pain Basic Data Set (ISCIPBDS) changes in pain prevalence, number of pain problems, and pain interference scores.** The assessments were conducted at Baseline, Endpoint, and Follow-up.

| | Baseline | Endpoint | Follow-up |
|---|---|---|---|
| Have you had any pain during the last seven days including today? | Yes | Yes | Yes |
| How many different pain problems did you have? | 4 | 2 | 2 |
| Pain interference | | | |
| 1. In general, how much has pain interfered with your day-to-day activities in the last week? | 0 | 0 | 0 |
| 2. In general, how much has pain interfered with your overall mood in the last week? | 1 | 0 | 1 |
| 3. In general, how much has pain interfered with your ability to get a good night's sleep in the last week? | 0 | 0 | 0 |

**Table S11. Background EMG during single-session and longitudinal neurophysiological assessments.** Root mean square (RMS) background electromyography (EMG) of flexor carpi radialis (FCR) and extensor carpi radialis (ECR) muscles was calculated over the 50-ms period preceding each stimulus across two sessions for motor evoked potential (MEP) (motor cortex stimulation (MCS); 8 intensities × 5 trials per session) and posterior root muscle (PRM) reflex (epidural spinal cord stimulation (ESCS); 5 intensities × 3 trials per session) recordings. For the longitudinal MEP assessments, background EMG was quantified across six assessment timepoints (8 intensities × 5 trials per assessment timepoint). Mean background EMG RMS values were below 5 μV, confirming that the participant remained at rest throughout the neurophysiological assessments. Values are presented as mean±SD.

| Background EMG RMS (μV) | | | | | | | | | |
|---|---|---|---|---|---|---|---|---|---|
| Single-session MEP assessment | | | | | Single-session PRM assessment | | | | |
| Muscle | BCI-ESCS | | Tonic-ESCS | | Muscle | BCI-ESCS | | Tonic-ESCS | |
| | Pre | Post | Pre | Post | | Pre | Post | Pre | Post |
| FCR | 1.98±0.25 | 1.92±0.44 | 1.73±0.18 | 1.94±0.32 | FCR | 2.33±1.52 | 2.08±0.27 | 2.09±0.13 | 2.35±0.27 |
| ECR | 2.13±0.72 | 2.20±0.66 | 2.06±0.72 | 2.04±0.66 | ECR | 3.15±5.41 | 4.04±3.37 | 2.18±0.66 | 2.14±0. 17 |

| Longitudinal MEP assessment | | | | | | |
|---|---|---|---|---|---|---|
| Muscle | Baseline | Day 6 | Day 10 | Day 21 | Endpoint | Follow-up |
| FCR | 3.43±1.43 | 1.76±0.19 | 1.84±0.12 | 2.11±0.28 | 1.70±0.15 | 2.01±0.65 |
| ECR | 1.84±0.11 | 2.15±0.85 | 1.89±0.54 | 2.38±0.79 | 1.98±0.57 | 2.49±1.48 |
